# Weight change and the incidence of cardiovascular diseases in adults with normal weight, overweight and obesity without chronic diseases; emulating trials using electronic health records

**DOI:** 10.1101/2020.05.14.20102129

**Authors:** M. Katsoulis, BD Stavola, KD Ordaz, M. Gomes, A Lai, P Lagiou, G Wannamethee, K Tsilidis, RT Lumbers, S Denaxas, A Banerjee, CA Parisinos, R Batterham, C Langenberg, H Hemingway

**Affiliations:** Institute of Health Informatics, University College London, UK; Health Data Research UK London, University College London, UK; Great Ormond Street Institute of Child Health, University College London, UK; Medical Statistics Department, London School of Hygiene and Tropical Medicine; Institute of Epidemiology & Health, University College London, UK; Dept. of Hygiene, Epidemiology and Medical Statistics, School of Medicine, National and Kapodistrian University of Athens, Greece; Department of Epidemiology and Biostatistics, Imperial College London, London, UK; Department of Hygiene and Epidemiology, University of Ioannina School of Medicine, Ioannina, Greece; Bart’s Heart Centre, St. Bartholomew’s Hospital, London, UK; Department of Epidemiology, Harvard T.H. Chan School of Public Health, Boston, MA, USA; The Alan Turing Institute, London, United Kingdom; The National Institute for Health Research University College London Hospitals Biomedical Research Centre, University College London, London, United Kingdom; British Heart Foundation Research Accelerator, University College London, London, United Kingdom; Amrita Institute of Medical Sciences, Kochi, India; Centre for Obesity Research, University College London, London, United Kingdom; University College London Hospitals Bariatric Centre for Weight Management and Metabolic Surgery, London; National Institute of Health Research, University College London Hospitals Biomedical Research Centre, London, United Kingdom; MRC Epidemiology Unit, University of Cambridge, Cambridge, UK

## Abstract

**Background:** Cross sectional measures of body mass index (BMI) are associated with cardiovascular disease (CVD) incidence, but less is known about whether weight change affects the risk of CVD.

**Methods:** We estimated the effect of 2-year weight change interventions on 7-year risk of CVD, by emulating hypothetical target trials using electronic health records. We identified 138.567 individuals in England between 1998 and 2016, aged 45-69 years old, free of chronic diseases at baseline. We performed pooled logistic regression, using inverse-probability weighting to adjust for baseline and time-varying variables. Each individual was classified into a weight loss, maintenance, or gain group.

**Findings:** In the normal weight, both weight loss and gain were associated with increased risk for CVD [HR vs weight maintenance=1.53 (1.18-1.98) and 1.43 (1.19-1.71 respectively)]. Among overweight individuals, both weight loss and gain groups, compared to weight maintenance, had a moderately higher risk of CVD [HR=1.20 (0.99–1.44) and 1.17 (0.99–1.38), respectively]. In the obese, weight loss had a lower risk lower risk of CHD [HR =0.66 (0.49–0.89)] and a moderately lower risk of CVD [HR =0.90 (0.72–1.13)]. When we assumed that a chronic disease occurred 1-3 years before the recorded date, estimates for weight loss and gain were attenuated among overweight individuals and estimates for weight loss were stronger among individuals with obesity.

**Interpretation:** Among individuals with obesity, the weight loss group had a lower risk of CHD and moderately lower risk of CVD. Weight gain increased the risk of CVD across BMI groups.

## INTRODUCTION

Lifestyle [1] and pharmacotherapy interventions [2,3] have been shown in randomised trials to be effective in achieving weight loss among individuals with overweight or obesity with high cardiovascular (CVD) risk because of type 2 diabetes, cardiovascular diseases or additional risk factors [3,4] In such high risk patients, there is evidence that weight loss is also beneficial for cardiovascular risk factors [4]. However, there are mixed findings on the effect of weight loss on CVD, as some studies found no effect of weight loss on fatal and non-fatal CVD [1,5,6], while a recent meta-analysis of trials reported moderate lower risk of CVD following weight loss [4]. In any case, there are no randomised controlled trials (RCTs) of any weight loss intervention assessing the effectiveness for primary prevention of cardiovascular diseases (CVDs) in otherwise healthy individuals with overweight or obesity. The prospect of performing such trials is low because of the very high number of participants required for reliable estimation of a primary preventive effect on CVD events. Therefore, it is not known whether weight loss reduces the risk of incident cardiovascular disease among people in the general population with overweight or obesity. This is particularly important as higher body mass index is associated with the onset of cardiovascular diseases [7] and the prevalence of obesity, already high, is predicted to risk further [8].

In such settings emulating target pragmatic trials using causal inference methods in large scale observational data may play a role in distinguishing effects of weight change in people with normal weight, overweight and obesity [9-12]. Observational studies of weight (or BMI) change are conflicting, some reporting increased risk of CVD [13-16], no association [17-20] or lower risk, especially after bariatric surgery in people with severe obesity [21]; weight gain has been associated with increased CVD risk in some studies[13-16], but not others[21]. Emulation of weight loss trials has been carried out in a consented cohort, the Nurses’ Health Study, which found no relationship between weight loss and CHD [17,18]. There have been no previous emulation of weight loss trials reporting separate effects within groups of people with normal weight, overweight or obesity. BMI at baseline is an important effect modifier in the relationship between weight change and CVD that is often overlooked in other observational studies as well [13,15]; weight loss might be more beneficial for individuals with obesity[21] than in the overweight[20], while weight loss might have adverse CVD in the normal weight[13].

In this study, we took advantage of the size, representativeness and contemporary measurements of weight, along with socioeconomic and demographic characteristics, available in Primary Care across England that are linked with hospital records and a national mortality registry [22]. We investigated hypothetical weight change interventions, applied for two years, and followed participants for an additional five years (i.e. 7 years follow-up in total) and assessed whether weight change affects the occurrence of CVDs. This was the largest study (∼138K) to estimate the effect of weight change on CVDs in individuals aged 45-69 years old and the first one to investigate this, separately in individuals with normal weight, overweight and obesity without chronic diseases.

## METHODS

### Data sources

We analysed data from individuals who were identified from the CALIBER programme in England between 1/1/1998 and 30/6/2016. CALIBER links anonymized coded electronic health records from three national data sources of patients across primary care (Clinical Practice Research Datalink), hospital care (Hospital Episode Statistics) and death registry (Office of National Statistics), from a large sample, representative of the population of England [22]. Methods for development of reproducible phenotypes and metadata have been described previously [23] and are available online (www.caliberresearch.org/portal). The study was approved by the Independent Scientific Advisory Committee (ISAC) of the Medicines and Healthcare products Regulatory Agency.

### Covariates

Height and weight measurements were based on primary care records and measured as part of routine care. We obtained all height and weight measurements available for the study population and calculated BMI as weight/ height^2^ (in kg/m^2^). In some cases, BMI was recorded, even if height or weight measurements were not. Our sample was divided into three groups based on standard clinical BMI cut-offs: individuals with i. normal weight (≥18.5 & <25 kg/m^2^), ii. overweight (≥25 & <30 kg/ m^2^) and iii. obesity (≥30 & <40 kg/m^2^).

We also used information on the following variables; age at baseline, sex, region, index of multiple deprivation (IMD), ethnicity, smoking status, physical activity, diabetes, cancer (apart from non-melanoma skin cancer), dementia, severe mental diseases (acute stress, phobia, anxiety, depression, schizophrenia, bipolar disorder and affective disorder), chronic kidney disease, chronic obstructive pulmonary disease, HIV, major inflammatory diseases (systemic lupus erythematosus, rheumatoid arthritis, gout, and inflammatory bowel disease), Parkinson’s disease, multiple sclerosis and renal failure (or initiation of dialysis). prevalence of hypertension, antihypertensive medication, number of weight measurements and number of clinical consultations every year.

### Target trials specification and emulation

We used electronic health records (EHR) to mimic target trials of weight change and cardiovascular outcomes in the English population. We first explicitly specified the target trials of interest and then used our observational data to emulate them [9-12]. We present the specification of the protocol of the target trial, along with the emulation of this protocol using EHR data in table 1 and we explain this procedure in detail in the Appendix.

**Table 1:**
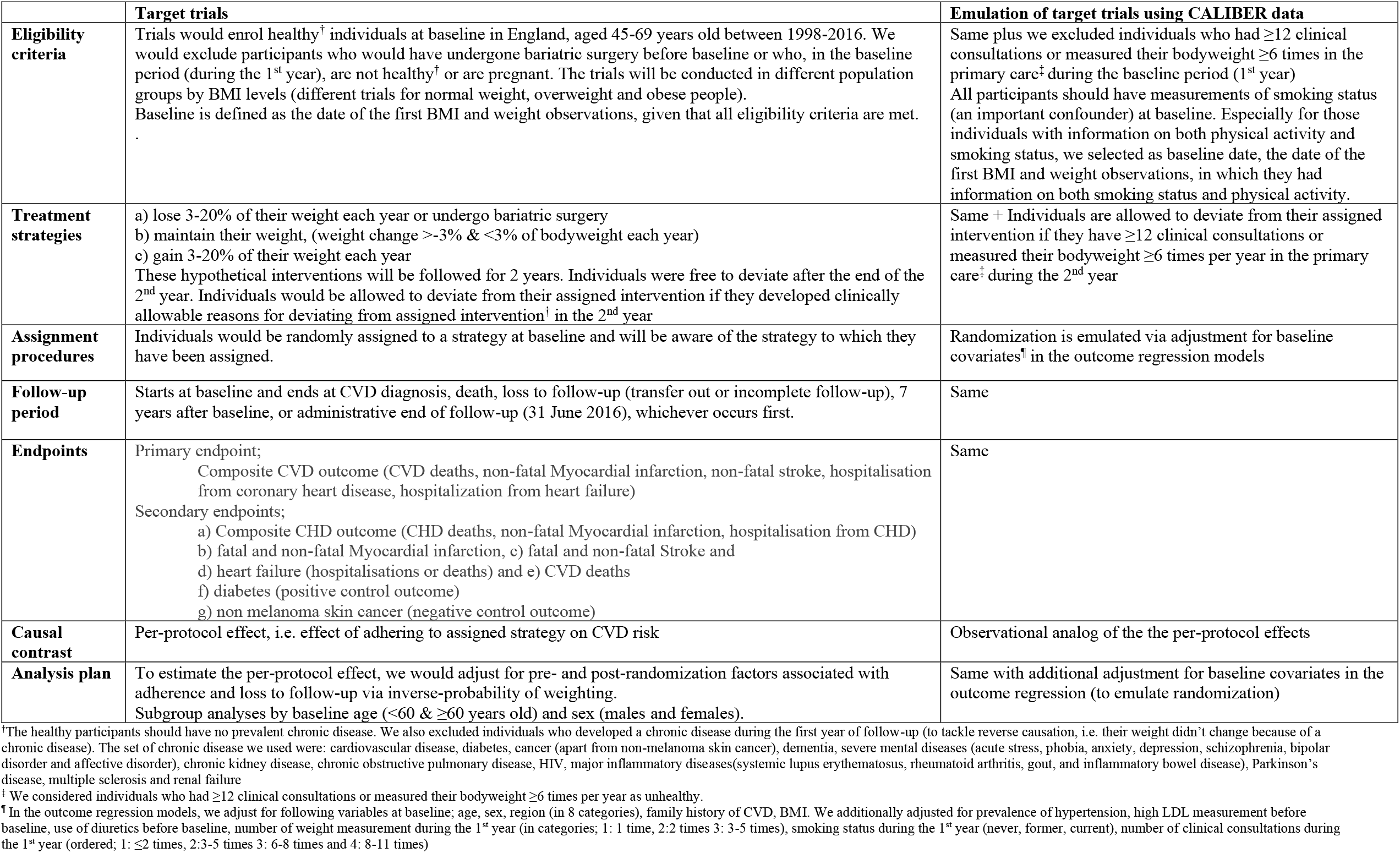
Specification and emulation of target trials to estimate the effect of weight change on various CVD outcomes. Description for the 3 main trials in normal weight, overweight and obese individuals

|  | Target trials | Emulation of target trials using CALIBER data |
| --- | --- | --- |
| **Eligibility criteria** | Trials would enrol healthy<sup>†</sup> individuals at baseline in England, aged 45-69 years old between 1998-2016. We would exclude participants who would have undergone bariatric surgery before baseline or who, in the baseline period (during the 1<sup>st</sup> year), are not healthy<sup>†</sup> or are pregnant. The trials will be conducted in different population groups by BMI levels (different trials for normal weight, overweight and obese people). Baseline is defined as the date of the first BMI and weight observations, given that all eligibility criteria are met.<br>. | Same plus we excluded individuals who had $\geq 12$ clinical consultations or measured their bodyweight $\geq 6$ times in the primary care<sup>‡</sup> during the baseline period (1<sup>st</sup> year) All participants should have measurements of smoking status (an important confounder) at baseline. Especially for those individuals with information on both physical activity and smoking status, we selected as baseline date, the date of the first BMI and weight observations, in which they had information on both smoking status and physical activity. |
| **Treatment strategies** | a) lose 3-20% of their weight each year or undergo bariatric surgery<br>b) maintain their weight, (weight change $>-3\%$ & $<3\%$ of bodyweight each year)<br>c) gain 3-20% of their weight each year<br>These hypothetical interventions will be followed for 2 years. Individuals were free to deviate after the end of the 2<sup>nd</sup> year. Individuals would be allowed to deviate from their assigned intervention if they developed clinically allowable reasons for deviating from assigned intervention<sup>†</sup> in the 2<sup>nd</sup> year | Same + Individuals are allowed to deviate from their assigned intervention if they have $\geq 12$ clinical consultations or measured their bodyweight $\geq 6$ times per year in the primary care<sup>‡</sup> during the 2<sup>nd</sup> year |
| **Assignment procedures** | Individuals would be randomly assigned to a strategy at baseline and will be aware of the strategy to which they have been assigned. | Randomization is emulated via adjustment for baseline covariates<sup>¶</sup> in the outcome regression models |
| **Follow-up period** | Starts at baseline and ends at CVD diagnosis, death, loss to follow-up (transfer out or incomplete follow-up), 7 years after baseline, or administrative end of follow-up (31 June 2016), whichever occurs first. | Same |
| **Endpoints** | Primary endpoint;<br>Composite CVD outcome (CVD deaths, non-fatal Myocardial infarction, non-fatal stroke, hospitalisation from coronary heart disease, hospitalization from heart failure)<br>Secondary endpoints;<br>a) Composite CHD outcome (CHD deaths, non-fatal Myocardial infarction, hospitalisation from CHD)<br>b) fatal and non-fatal Myocardial infarction, c) fatal and non-fatal Stroke and<br>d) heart failure (hospitalisations or deaths) and e) CVD deaths<br>f) diabetes (positive control outcome)<br>g) non melanoma skin cancer (negative control outcome) | Same |
| **Causal contrast** | Per-protocol effect, i.e. effect of adhering to assigned strategy on CVD risk | Observational analog of the the per-protocol effects |
| **Analysis plan** | To estimate the per-protocol effect, we would adjust for pre- and post-randomization factors associated with adherence and loss to follow-up via inverse-probability of weighting.<br>Subgroup analyses by baseline age ($<60$ & $\geq 60$ years old) and sex (males and females). | Same with additional adjustment for baseline covariates in the outcome regression (to emulate randomization) |

<sup>†</sup>The healthy participants should have no prevalent chronic disease. We also excluded individuals who developed a chronic disease during the first year of follow-up (to tackle reverse causation, i.e. their weight didn't change because of a chronic disease). The set of chronic disease we used were: cardiovascular disease, diabetes, cancer (apart from non-melanoma skin cancer), dementia, severe mental diseases (acute stress, phobia, anxiety, depression, schizophrenia, bipolar disorder and affective disorder), chronic kidney disease, chronic obstructive pulmonary disease, HIV, major inflammatory diseases(systemic lupus erythematosus, rheumatoid arthritis, gout, and inflammatory bowel disease), Parkinson's disease, multiple sclerosis and renal failure
<sup>‡</sup> We considered individuals who had $\geq 12$ clinical consultations or measured their bodyweight $\geq 6$ times per year as unhealthy.
<sup>¶</sup> In the outcome regression models, we adjust for following variables at baseline; age, sex, region (in 8 categories), family history of CVD, BMI. We additionally adjusted for prevalence of hypertension, high LDL measurement before baseline, use of diuretics before baseline, number of weight measurement during the 1<sup>st</sup> year (in categories; 1: 1 time, 2:2 times 3: 3-5 times), smoking status during the 1<sup>st</sup> year (never, former, current), number of clinical consultations during the 1<sup>st</sup> year (ordered; 1: $\leq 2$ times, 2:3-5 times 3: 6-8 times and 4: 8-11 times)

### Statistical analysis of the emulated trials

The analysis involved several components linked to the trial emulation steps, as outlined below.

#### i. Data structure

Follow-up was divided into one-year periods in which the weight change intervention per year was recorded, along with information on confounders, CVD outcome (1:yes, 0:no), death (1:yes, 0:no) and loss to follow-up (1:yes, 0:no) recorded during each year. We classified each individual into one of the following groups, corresponding to their observed weight trajectories during the first year of follow-up: (1) weight loss (−20% to −3% change in bodyweight per year or bariatric surgery), (2) weight maintenance (−3% to +3% change in bodyweight per year), or (3) weight gain (+3% to +20% change in bodyweight per year)

#### ii. Outcome regression

Pooled logistic regression model was used to estimate the hazard ratios of the hypothetical interventions and the cumulative incidence risk curves of each intervention[9].

#### iii. Emulating randomisation at baseline

Time of entry in the emulated trials was considered the date of the first BMI and weight observations, when all eligibility criteria were met (see table 2 and Appendix, section 1). In our case, it is important to mention that each time point corresponds to one-year duration of our (hypothetical) interventions. For this reason, to emulate randomisation at baseline, we adjusted for: age (in years), sex (men/ women), BMI (in kg/m^2^), prevalence of hypertension (before baseline; yes/no), record of high LDL levels (before baseline; yes/no), use of diuretics (before baseline; yes/no); family history of CVD (yes/no); hypertension (during the 1st year, yes/no); high LDL levels (during the 1st year; yes/no), use of diuretics (during the 1st year); smoking status (during the 1st year; three categories: never, former and current), number of weight measurements (during the 1st year; three categories: 1, 2 and 3-5 meas.), number of clinical consultations (during the 1st year; four categories: 1-2, 3-5, 6-8 and 9-11 consultations) and region (in categories; London, South West, South Central, South East, East, West Midlands, Central North and North West). This means that we are assuming randomisation was conditional on these baseline covariates (equivalent to no unobserved confounding given these).

**Table 2:**
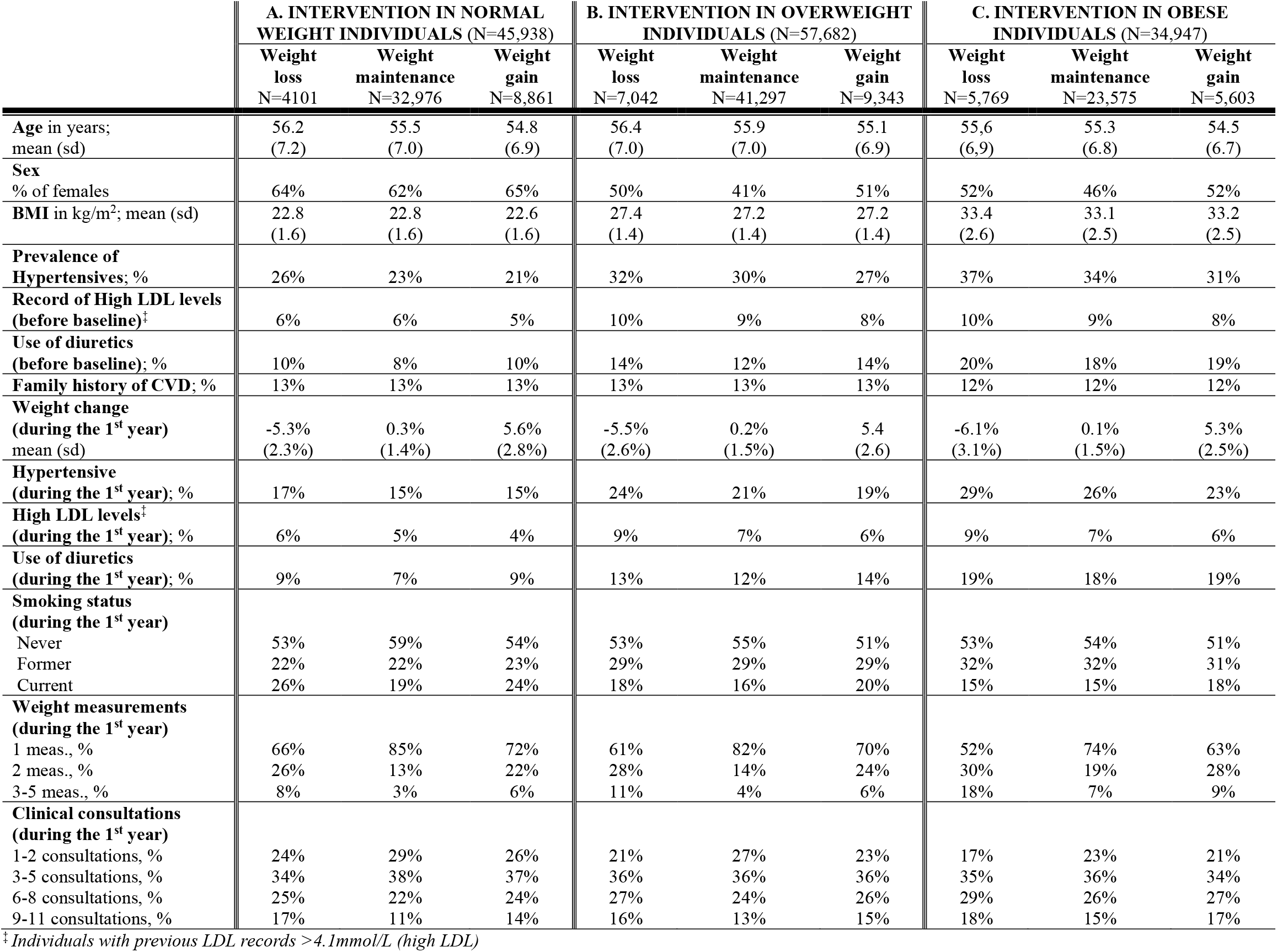
Characteristics of individuals at baseline and during the 1^st^ year, by BMI group and hypothetical weight change intervention

#### iv. Dealing with non-adherence to intervention

Non-adherence occurred when individuals were allocated (i.e. observed) to a particular weight change group in the first year but deviated from this group in second. Inverse probability of treatment weights (IPTW) were used to adjust for time-fixed and time-dependent confounders[9,12]. In IPTW, we weight each observation by the inverse of the probability of an individual having received his/her observed weight change intervention during the 2^nd^ year, given his/her past intervention and prognostic factors history. To calculate the denominator of the IPTW, we used multinomial logistic regression models for the observed weight change intervention (i.e. weight loss, maintenance and gain) during the 2^nd^ year, in which we included the determinants of the observed intervention at the 2^nd^ year, i.e. prognostic factors measured before baseline, during the 1^st^ year (as described above) and during the 2^nd^ year of these interventions, along with the observed weight change intervention during the 1^st^ year. The IPTW remained constant after the second year, because we were interested in the effect of a weight change intervention sustained over 2 years only. After calculating the IPTW for the received intervention in the second year, individuals were then censored during the 2^nd^ year, if they deviate from their assigned intervention. For those individuals who develop a chronic disease other than CVD during the 2^nd^ year (and thus were free to deviate in the 2^nd^ year as well from their assigned intervention), they were assigned the weight of 1 across all time points. We remark that we used the non-stabilised weights, because the regime of the trials was dynamic [12] (i.e. we had specified clinically allowable reasons after which individuals were free to deviate from their initial intervention)

#### v. Dealing with loss to follow-up

Additional adjustment for pre- and post-randomisation prognostic factors of loss to follow-up was also used through inverse probability of censoring weighting (IPCW), to estimate the effect of the interventions, had the participants remained uncensored during the follow-up[12]. These analyses were thus valid under missing at random, given the covariates used to model the censoring mechanism. For more information, see Appendix (Section 3).

#### vi Final calculation of IP weights

IPTW and IPCW were multiplied at each time point. The final weight for each individual for a specific time was taken as the product of his/her weights up until that time point. We truncated weights >15 (which were higher than the 99^th^ percentile of weights) and set it to 15.

#### vii. Risk curves

We additionally estimated absolute risks for CVD, CHD and diabetes by fitting the pooled logistic models that were mentioned above, including product terms between treatment and follow-up time (time, squared time and cubic time) to allow for time varying effects. The estimated parameters were then used to calculate the cumulative incidence of CVD, CHD and diabetes (see details in the Appendix – section 3).

#### viii. Variance estimators

We used robust variance estimators to calculate 95% CI for the hazard ratio estimates, and we used non-parametric bootstrapping from 500 samples to obtain percentile-based 95% CI for the cumulative incidence estimates. For more details, see Appendix (Sections 2 and 3)

### Positive and negative control outcomes

In the analysis of the positive control outcome, we expect to observe a (well established from the literature) non-null relationship between the exposure and the outcome, while in the analysis of the negative control outcome we anticipate to estimate no association, as there in no link between the exposure and the outcome. We used positive and negative control outcomes, because any deviation from the expected associations would help us detect potential biases due to unmeasured confounding in our emulated trials. We applied the hypothetical interventions as described in table 1 and the Appendix (Section 2), using as endpoints a) diabetes (positive control outcome) and b) non-melanoma skin cancer (negative control outcome). We chose to use diabetes as a positive control outcome, as we expected that compared to weight maintenance, weight loss would be related to lower diabetes risk and weight gain to higher diabetes risk [24]. We additionally chose non-melanoma skin cancer as a negative control outcome, because there is no established connection between weight change and non-melanoma skin cancer.

### Sensitivity analysis

We applied the eligibility criteria to individuals in the CALIBER database, plus we required that the individuals have measurements of index of multiple deprivation (IMD), ethnicity and physical activity (assuming that last observation carries forward for at most four years). We took these variables into account when adjusting the pooled logistic regression (to emulate randomisation) and when estimating the IP weights. We also applied subgroup analysis by age and sex.

Moreover, to take into consideration potential pre-clinical diseases, we assumed that a chronic disease occurred one, two or three years before it was recorded during the follow-up and we assessed whether our estimates were robust to this decision. The set of chronic diseases considered for this sensitivity analysis was: diabetes, cancer (apart from non-melanoma skin cancer), dementia, severe mental diseases (acute stress, phobia, anxiety, depression, schizophrenia, bipolar disorder and affective disorder), chronic kidney disease, chronic obstructive pulmonary disease, HIV infection, major inflammatory diseases (systemic lupus erythematosus, rheumatoid arthritis, gout, and inflammatory bowel disease), Parkinson’s disease, multiple sclerosis and renal failure (or initiation of dialysis).

## RESULTS

Of 1,161,264 individuals, aged 45-69, in the primary care database with BMI and weight measurements between 1/1/1998 and 30/6/2016, 138,567 were eligible for the hypothetical trials conducted separately in individuals with normal weight, overweight and obesity (Figure 1). Specifically, the emulated trials for normal weight, overweight and people with obesity included 45,938, 57,682 and 34,947 individuals, respectively. Additionally, we observed that the percentage of individuals who adhered to their “assigned” intervention during the 2^nd^ year was much higher in the weight maintenance group in all 3 trials (72%, 72% and 67% in individuals with normal weight, overweight with obesity respectively), compared to the other arms. Individuals across intervention groups were similar with respect to their baseline characteristics (Table 2). In the weight maintenance group, the percentage of women was lower compared to the other two groups. Additionally, individuals in the weight maintenance group had fewer clinical consultations and had their weight measured within primary care less frequently during the first year, compared with those in the other two hypothetical interventions.

**Figure 1:**
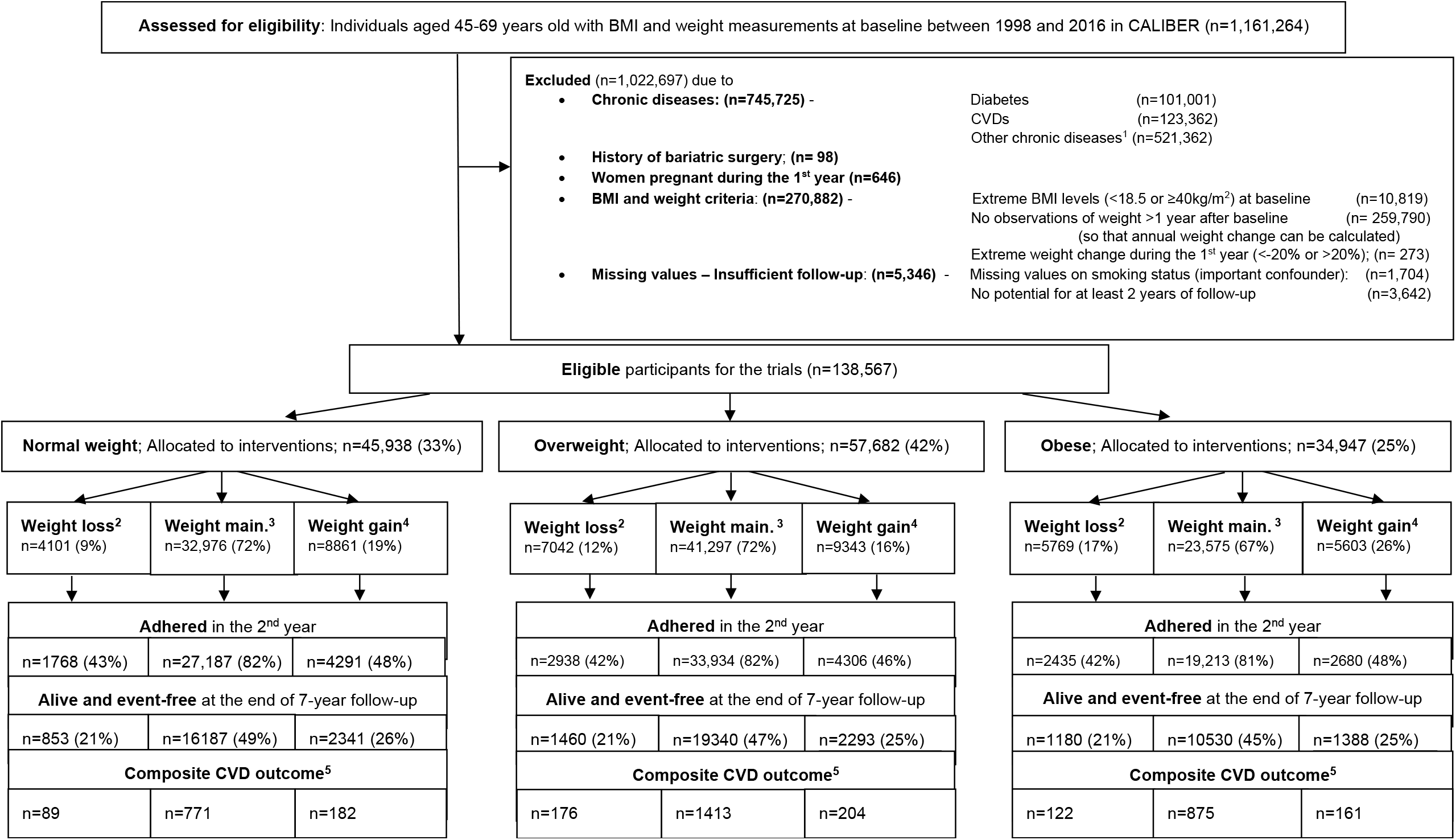
CONSORT diagram - Emulating 2-year weight change interventions in normal weight, overweight and obese individuals ^1^The set of chronic diseases, which occurred before baseline or during the 1^st^ year of follow-up, consists of; CVD, diabetes, cancer (apart from non-melanoma skin cancer), mental health diseases (acute stress, phobia, anxiety depression, schizophrenia, bipolar disorder and affective disorder), chronic kidney disease, major inflammatory diseases (systemic lupus erythematosus, rheumatoid arthritis, gout and ulcerative colitis), Parkinson’s disease, multiple sclerosis and renal failure. We also considered that those with ≥6 weight measurement and ≥12 clinical consultations per year in their practice would have a chronic disease ^2^Weight loss arm: lose >3% & <20% of bodyweight each year or undergo bariatric surgery ^3^Weight maintenance arm: weight change ≥-3% & ≤3% of bodyweight each year ^4^Weight gain arm: gain >3% & <20% of bodyweight each year. ^5^Incident CVDs – primary outcome consisted of CVD deaths, non-fatal Myocardial infarction, non-fatal stroke, hospitalisation from coronary heart disease

### Diabetes and non-melanoma skin cancer as positive and negative control outcomes

In analyses of diabetes as a positive control outcome, the cumulative risk was higher in the weight loss group than in weight maintenance group during the first 2-3 years, in all BMI groups (see figure 2). The corresponding hazard ratios of weight loss (vs weight maintenance) were, across BMI groups, greater than 1 up to the first 3 years [HR=1.39 (0.96, 2.00), HR=1.53 (1.27, 1.93) and HR=1.32 (1.13, 1.54) in the normal weight, overweight and individuals with obesity respectively], and lower than 1 after the 3^rd^ year [HR=0.76 (0.38, 1.53), HR=0.88 (0.60, 1.28) and HR=0.64 (0.46, 0.88) in the normal weight, overweight and individuals with obesity respectively]. In analyses of non-melanoma skin cancer as a negative outcome control, the hazard ratios of both weight gain and weight loss were close to one (Figure S1).

**Figure 2:**
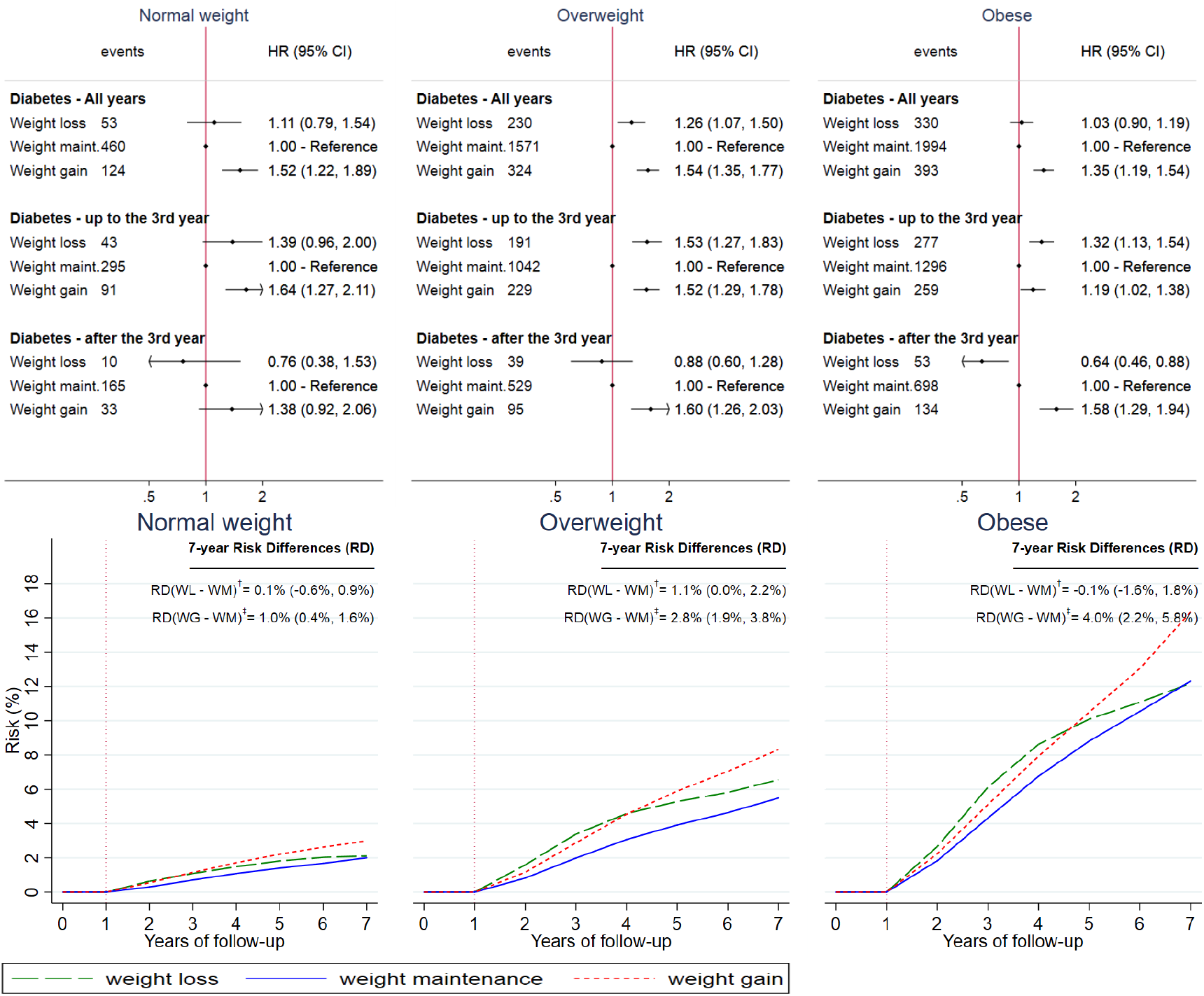
Estimated hazard ratios (upper panel) and cumulative incidence curves^¶^ (lower panel) for diabetes (i.e. positive control outcome) under hypothetical weight change interventions, by BMI group ^¶^For the technical details for the estimation of the cumulative risk curves, see Appendix (Section XXX) ^†^RD(WL - WM): Risk Difference (Weight loss intervention - Weight maintenance intervention) ^‡^RD(WG - WM): Risk Difference (Weight gain intervention - Weight maintenance intervention) Individual were free of diabetes during the 1^st^ year of the intervention (see vertical dotted line at year 1)

### Occurrence of chronic diseases (other than CVD) during follow-up

In all the emulated trials, we found that the incidence of chronic diseases during the first four years was higher in the weight loss and weight gain arm, compared to the weight maintenance arm, across BMI groups (Figure S2). More specifically, during the 2^nd^ year, the incidence of chronic diseases was 24% in the normal weight, 21% in the overweight and 23% in people with obesity in the weight loss arm. The corresponding incidence was 7% (normal weight), 8% (overweight) and 10% (obese) in the weight maintenance group and 13% (normal weight), 18% (overweight) and 20% (obese) in the weight gain arm. The same pattern was observed in the 3^rd^ and 4^th^ year as well.

### Emulated trials in the normal weight

Among normal weight individuals, those in the weight loss and weight gain groups had a higher risk for the composite CVD outcome (Figure 2). Compared to weight maintenance, the hazard ratios for weight loss were 1.53 (1.18 – 1.98) and for weight gain 1.43 (1.19 - 1.71) (Figure 3). A similar pattern was observed for most of the secondary outcomes. Of note, the most pronounced increased risks for weight loss was for heart failure [figure 4, HR=2.72 (1.54-4.81)] and CVD deaths [HR=2.55 (1.37-4.77)]. Estimates were similar in subgroups of individuals defined at baseline according to age, but different for females compared with males (HR= 1.72 (1.22-2.41) for men, HR=1.28 (0.84 −1.96) for women, comparing weight loss to weight maintenance) (see Appendix. Figure S3). In sensitivity analyses, estimates were similar with additional adjustment for Index of Multiple Deprivation, ethnicity or physical activity (see Appendix Figure S4). Estimates were also similar when we assumed that a set of chronic diseases occurred one, two or three years before they were recorded in the database (see Figure 3 and Appendix Figure S5).

**Figure 3:**
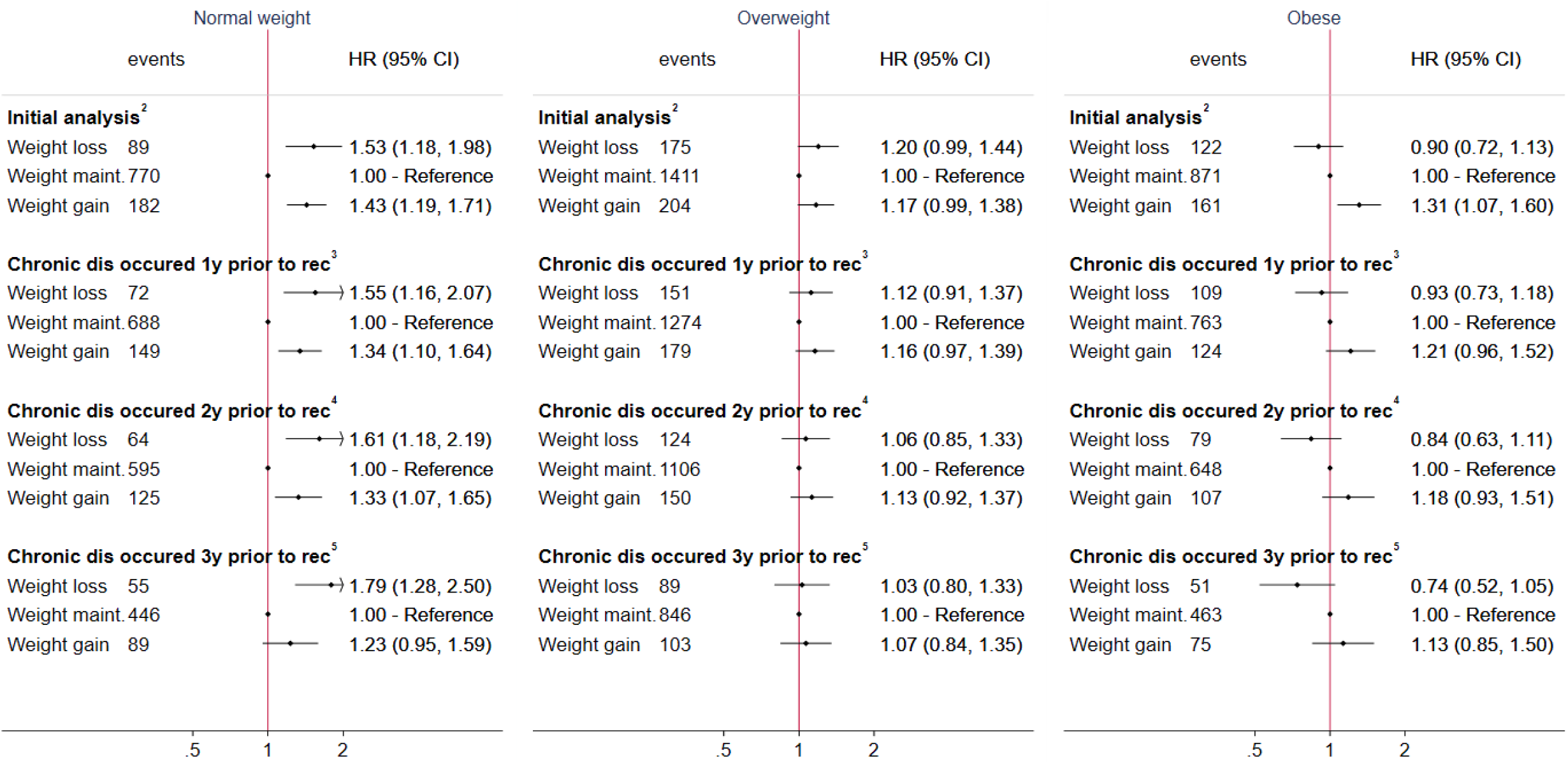
Estimated hazard ratios for cardiovascular diseases comparing hypothetical weight change interventions, by BMI group. Results from initial analysis as well from the sensitivity analysis, in which a set of chronic diseases^1^ was assumed to occur one, two or three years prior to the recorded date ^1^Set of chronic diseases consists of:, diabetes, cancer (apart from non-melanoma skin cancer), dementia, severe mental diseases (acute stress, phobia, anxiety, depression, schizophrenia, bipolar disorder and affective disorder), chronic kidney disease, chronic obstructive pulmonary disease, HIV, major inflammatory diseases(systemic lupus erythematosus, rheumatoid arthritis, gout, and inflammatory bowel disease), Parkinson’s disease, multiple sclerosis and renal failure ^2^Assuming that the set of chronic diseases occurred at the recorded date ^3^Assuming that the set of chronic diseases occurred one year prior to the recorded date ^4^Assuming that the set of chronic diseases occurred two years prior to the recorded date ^5^Assuming that the set of chronic diseases occurred three years prior to the recorded date

**Figure 4:**
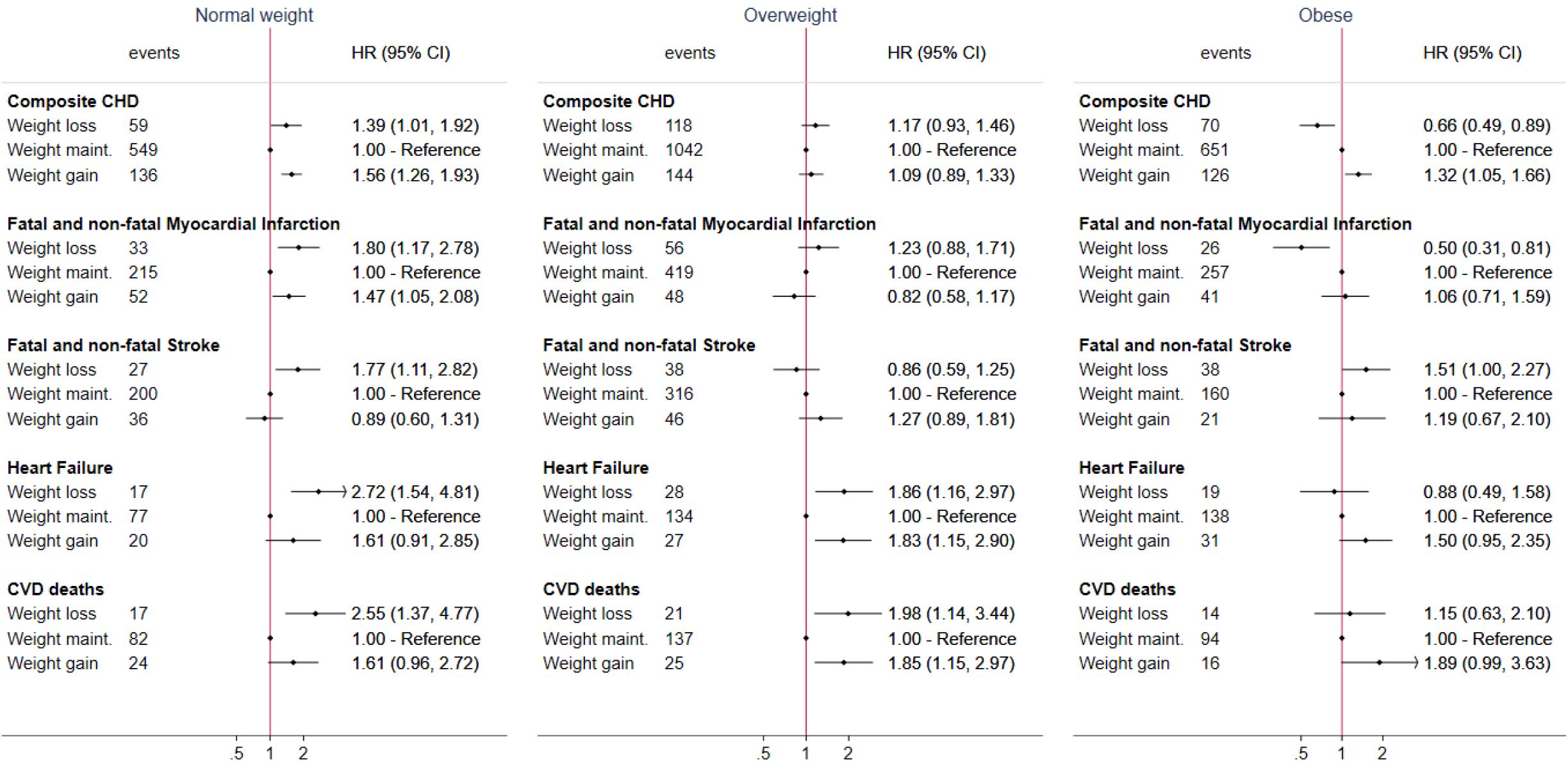
Estimated hazard ratios for cardiovascular diseases (secondary outcomes) comparing hypothetical weight change interventions, using pooled logistic regression

### Emulated trials in the overweight

Among individuals with overweight, those in the weight loss and weight gain groups had a higher risk for CVD, compared to those in the weight maintenance groups (HR= 1.20 (0.99 – 1.44) for weight loss and 1.17 (0.99 – 1.38) for weight gain] (Figure 3). Estimates were similar with additional adjustment for Index of Multiple Deprivation, ethnicity or physical activity (see Appendix Figures S4) and in subgroup analyses by age and sex (see Appendix Figure S6). However, we did observe some differences, when considering that chronic diseases occurred one, two or three years before the recorded diagnosis date. Estimates for weight loss and weight gain (vs weight maintenance) were attenuated with increasing lags for chronic disease records [Figure 3, Figure S5 (Appendix)], which was particularly pronounced for weight loss (HR: 1.12 (0.91–1.36), 1.06 (0.85–1.33) and 1.03 (0.80–1.33), under lags of one, two, and three years, respectively).

### Emulated trials in individuals with obesity

Among individuals with obesity, those in the weight gain group had a higher risk for CVD [HR =1.31 (1.07-1.60)], compared to those in the maintenance group (Figure 3). Individuals in the weight loss group had a moderately lower risk for CVDs [HR vs weight maintenance=0.90 (0.72-1.13)]. However, in the analysis of secondary outcomes, those in the weight loss group had a much lower risk of CHD [HR =0.66 (0.49-0.89)] and myocardial infarction [HR =0.50 (0.31-0,81)]. The estimated 7-year risk of CHD was 2.7% (2.0% to 3.6%), 4.2% (3.9% to 4.6%) and 5.4% (4.3% to 6.5%) for the weight loss, maintenance, and gain groups, respectively (Figure 5). On the contrary, individuals in the weight loss group had higher risk for stroke [Figure 4, HR=1.51 (1.00-2.27)]. Estimates were similar with additional adjustment for Index of Multiple Deprivation, ethnicity or physical activity (Appendix Figures S4). However, hazard ratios for CVD comparing weight loss to weight maintenance differed by sex [in Figure S7, HR= 1.03 (0.79-1.34) in men and HR=0.65 (0.43-1.01) in women] and age [HR= 0.83 (0.61-1.14) in individuals aged 45-59 and HR=1.03 (0.73-1.46) in individuals aged 60-69]. Moreover, estimated hazard ratios comparing weight loss to weight maintenance decreased with increasing lags for chronic disease records (HR= 0.93 (0.73 – 1.18), 0.84 (0.63 – 1.11) and 0.74 (0.52 – 1.05) under lags of one, two or three years, respectively) (Figure 3).

**Figure 5:**
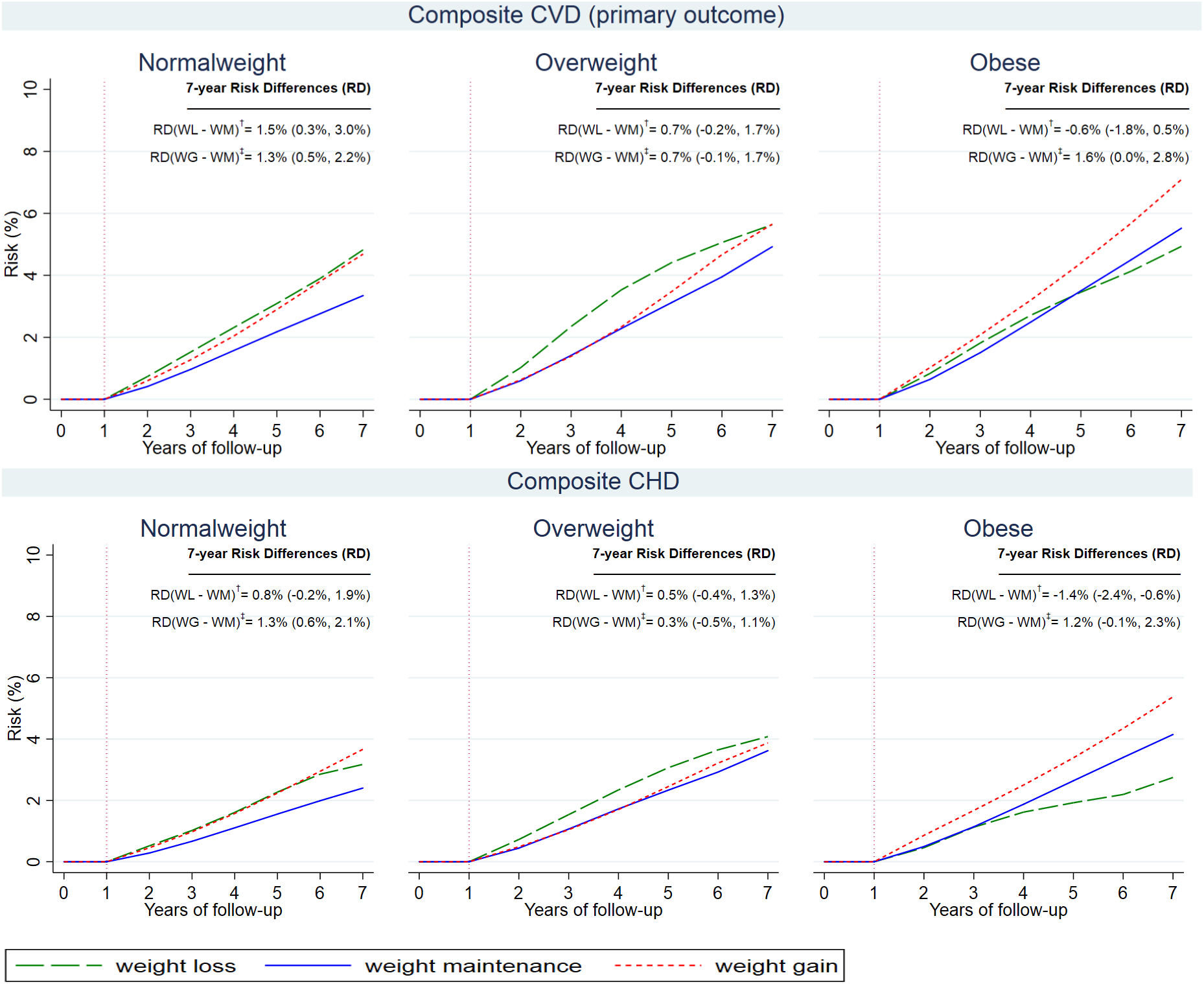
Cumulative incidence curves^¶^ for composite CVD and coronary diseases under hypothetical weight change interventions, by BMI group ^¶^For the technical details for the estimation of the cumulative risk curves, see Appendix (Section 3) ^†^RD(WL - WM): Risk Difference (Weight loss intervention - Weight maintenance intervention) ^‡^RD(WG - WM): Risk Difference (Weight gain intervention - Weight maintenance intervention) Individual were free of CVD during the 1^st^ year of the intervention (see vertical dotted line at year 1)

## DISCUSSION

In this study, we used large-scale EHR data to emulate target trials of weight loss, maintenance and gain interventions, estimating their effect on incident CVD, separately among individuals with normal weight, overweight, and obesity. We found that, among normal weight individuals, the weight maintenance group had a lower CVD risk, compared with the weight gain and the weight loss group. Among individuals with overweight, the weight loss and weight gain groups had a slightly higher CVD risk, compared with weight maintenance; however, this finding was not robust in sensitivity analyses for unmeasured confounding by preclinical diseases. Among individuals with obesity, the weight loss group had a lower risk of CHD and moderately lower of CVD, while the weight gain group had a higher CVD risk, compared with the weight maintenance group.

### Strengths and limitations

Key strengths of this study include the use of large-scale, linked-electronic health records and the application of cutting-edge causal inference methods [9-12], the combination of which gave us the opportunity to emulate weight change trials in 138,567 otherwise healthy individuals, who were followed for up to seven years. These target trials would be impossible to conduct, as most of the real-world trials usually recruit people with chronic disease and do not follow them up for sufficient time to measure “hard” outcomes, like CVD [3]. The large sample size gave as a unique opportunity to investigate the relationship between weight change and CVD among different population groups, by BMI group, as well as by age and sex. Individuals with normal weight, overweight and obesity have different underlying risk for CVD[7], hence it is more appropriate to study them separately. Most observational studies cannot focus on this relationship in this resolution, mainly due to restrictions posed by their sample size. Another strength of this study was the utilisation of clinically relevant measurements of weight and height, along with records for various comorbidities in the EHR data.

Several limitations should be noted. The observational study design of this research, despite our use of causal inference methods is still prone to unmeasured confounding. We tried to observe to what extent the bias due to preclinical diseases would affect our results by emulating the same interventions using positive and negative control outcomes. We observed that weight loss was related to higher risk for diabetes, compared to weight maintenance, during the first 2-3 years. This unexpected result may be due to unmeasured confounding by subclinical disease (i.e. reverse causation). We also noticed this potential violation of “recruiting” healthy individuals, when we observed that the percentage of individuals who developed a chronic disease during the 2^nd^ year was much higher in the weight loss compared to the weight maintenance intervention. To address this problem, we considered that the actual date of the occurrence of a chronic condition was one, two or three years before the recorded date and observed whether there was any specific trend in our findings from this sensitivity analysis. Moreover, it is important to mention that under these hypothetical interventions, we could not identify the way different people lost, maintained or gained weight (i.e. whether it was through lower caloric intake or higher levels of physical activity or because of a non-recorded disease). Different methods of modifying bodyweight may have different effects on CVD risk [25,26]. Moreover, we utilised EHR data, so we had to make additional assumptions in comparison to other studies that used cohort data[17,18]. These included the assumptions that a) the last observation carried forward for at most 4 years for smoking status (and physical activity in the sensitivity analysis) and b) there was a linear trend for weight change between 2 bodyweight measurements recorded less than 4 years apart. The assumption for smoking status is more likely to hold, as most people do not change lifestyle habits very frequently. Moreover, we were unable to account for different patterns of weight change over the course of a year that may lead to the same measure of annual weight change. Finally, we could not identify whether weight change was intentional or not in our study. It is plausible to assume unintentional weight change, especially for those interventions that are not clinically relevant (i.e. weight loss in the normal weight).

### Meaning of the study: possible explanations and implications

First, we observed that weight maintenance, compared to weight loss or gain, was linked with lower risk for all the CVD endpoints, in normal weight individuals. These results were in line with the cohort studies that have shown that normal weight individuals have less risk for developing CVD [7] compared to the other BMI group and thus infer that these individuals should not gain or lose weight.

Among individuals with overweight, we initially found evidence that both weight gain and loss may moderately increase CVD risk; however, estimates were attenuated (especially for weight loss) in sensitivity analyses when we assumed that a chronic disease might have occurred before the recorded date. These findings are similar to those of two previous studies [18,19]. Danaei et al[18] previously emulated a target trial of weight loss and CHD and found no relationship among participants with BMI≥25kg/m^2^ (overweight and obese together). In the 2^nd^ study by Wannamethee et al, which was not within the trial emulation framework, the authors’ inability to capture a protective association might have been due to the moderate sample size (∼3K overweight individuals among whom only ∼400 lost weight [19].

In people with obesity, weight gain was related to a higher risk for CVD, which is in line with the numerous reports that have linked high BMI to the onset of CVD [7]. Weight loss was not related to a clear CVD risk reduction, even though in the sensitivity analysis we observed that the more years we assumed the set of chronic diseases happened before the recorded date, the lower the hazard ratio for CVD of the weight loss was. When we stratified the analysis by age and sex, we found that lower risk of CVD for women and younger individuals. Furthermore, weight loss was related to lower risk for CHD and especially to fatal and non-fatal myocardial infarction. Individuals with obesity should reduce their bodyweight to lower their risk for coronary heart diseases. US Preventive Services Task Force currently recommends behavioural weight loss interventions for individuals with obesity (BMI ≥30 kg/m^2^)[27]. Nevertheless, the expected beneficial effect from weight reduction was only detectable for CHD, but not for stroke in our study. The effect of weight loss might not be homogeneous across pathologically diverse CVD outcomes. Although the American Stroke Association recommends weight loss for people with overweight or obesity for the primary prevention of ischaemic stroke [28] we did not observe any benefit with weight reduction. This may be due to the fact that BMI is an imprecise measure of body fat and does not distinguish between fat and muscle mass. We were not able to measure central adiposity which has been shown to be the adiposity measure more associated with stroke risk than BMI [29], as weight loss may reflect loss in muscle mass [18].

### Unanswered questions and future research

Our study could not clarify how individuals lost, maintained or gained weight. This unanswered question needs to be addressed by future research in the field. More studies should focus on how to emulate trials from observational data to investigate how adherence to different diets or physical activity levels that result in weight change, could affect a range of CVD outcomes. Finally, future studies in different population groups, stratified by age, sex and BMI group should be undertaken to maximize the benefits of stratified approaches to cardiovascular health.

## Conclusions

Weight maintenance had the lowest risk for CVD among individuals with normal weight. Among individuals with obesity, the weight loss group had a lower risk of CHD and moderately lower risk of CVD. Weight gain increased the risk of CVD across BMI groups. Our results may help to inform policy guidelines for cardiovascular prevention.

## Data Availability

N/A

## ACKNOWLEDGEMENTS

We would like to thank Prof Miguel Hernan and Dr Barbra Dickerman for the discussions and their useful suggestions during the drafting of the paper.

This study was approved by the Medicines and Healthcare Products Regulatory Agency Independent Scientific Advisory Committee protocol references: 18_010. This study is based in part on data from the Clinical Practice Research Datalink obtained under license from the UK Medicines and Healthcare products Regulatory Agency. The data are provided by patients and collected by the NHS as part of their care and support. The interpretation and conclusions contained in this study are those of the author(s) alone. Hospital Episode Statistics Copyright (2019) is reused with the permission of The Health & Social Care Information Centre. All rights reserved. The Office of Population Censuses and Surveys Classification of Interventions and Procedures, codes, terms and text is Crown copyright (2016) published by Health and Social Care Information Centre, also known as NHS Digital and licensed under the Open Government Licence (available at www.nationalarchives.gov.uk/doc/open-government-licence/open-government-licence.htm).

This study was carried out as part of the CALIBER programme (https://www.ucl.ac.uk/health-informatics/caliber). CALIBER, led from the UCL Institute of Health Informatics, is a research resource consisting of linked electronic health records phenotypes, methods and tools, specialized infrastructure, and training and support. The views expressed are those of the author(s) and not necessarily those of the National Health Service, the National Institute for Health Research, or the Department of Health. This paper represents independent research [part] funded by the National Institute for Health Research (NIHR) Biomedical Research Centre at UCL Hospitals. The views expressed are those of the author(s) and not necessarily those of the NHS, the NIHR or the Department of Health and Social Care.

## CONTRIBUTORS

MK designed the study, did the literature search, conducted the statistical analysis and interpreted the results, and wrote the manuscript. BDS designed the study, interpreted the results, and commented on the manuscript. KDO designed the study, interpreted the results, and commented on the manuscript. MG interpreted the results and commented on the manuscript. RB interpreted the results and commented on the manuscript. AGL, PL, GW, KK, GW, RTL, SD, AB, CAP, CL, RB, commented on the manuscript. HHH designed the study, interpreted the results, and commented on the manuscript

## REFERENCES

1. Rock CL, Flatt SW, Pakiz B, Taylor KS, Leone AF, Brelje K, Heath DD, Quintana EL, Sherwood NE. Weight loss, glycemic control, and cardiovascular disease risk factors in response to differential diet composition in a weight loss program in type 2 diabetes: a randomized controlled trial. Diabetes Care. 2014;37(6):1573–80.

2. Bohula EA, Wiviott SD, McGuire DK, et al.Cardiovascular Safety of Lorcaserin in Overweight or Obese Patients.N Engl J Med. 2018 Sep 20;379(12):1107–1117

3. Khera R, Pandey A, Chandar AK, Murad MH, Prokop LJ, Neeland IJ, Berry JD, Camilleri M, Singh S.Effects of Weight-Loss Medications on Cardiometabolic Risk Profiles: A Systematic Review and Network Meta-analysis. Gastroenterology. 2018;154(5):1309-1319.e7.

4. Ma C, Avenell A, Bolland M, et al. Effects of weight loss interventions for adults who are obese on mortality, cardiovascular disease, and cancer: systematic review and meta-analysis. BMJ. 2017; 359: 4849

5. Ackermann RT, Liss DT, Finch EA, et al. A Randomized Comparative Effectiveness Trial for Preventing Type 2 Diabetes. Am J Public Health 2015;359:2328–34

6. Look AHEAD Research Group. Cardiovascular effects of intensive lifestyle intervention in type 2 diabetes. N Engl J Med 2013;359:145–54

7. Mongraw-Chaffin ML, Peters SA, Huxley RR, Woodward M. The sex-specific association between BMI and coronary heart disease: a systematic review and meta-analysis of 95 cohorts with 1.2 million participants. Lancet Diabetes Endocrinol. 2015;3(6):437–49

8. Ward ZJ, Bleich SN, Cradock AL, Barrett JL, Giles CM, Flax C, Long MW, Gortmaker SL.Projected U.S. State-Level Prevalence of Adult Obesity and Severe Obesity. N Engl J Med. 2019;381(25):2440–2450

9. Dickerman BA, García-Albéniz X2, Logan RW, Denaxas S, Hernán MA.Avoidable flaws in observational analyses: an application to statins and cancer.Nat Med. 2019;25(10):1601–1606

10. Hernán MA, Robins JM. Using big data to emulate a target trial when a randomized trial is not available. American Journal of Epidemiology 2016; 183(8):758–764.

11. Danaei G, LA Garcia-Rodriguezc, Fernandez-Cantero O, Logan R, Hernan MA. Electronic medical records can be used to emulate target trials of sustained treatment strategies.J Clin Epidemiol. 2018; 96: 12–22.

12. Hernán MA, Robins JM (2020). Causal Inference: What If. Boca Raton: Chapman & Hall/CRC

13. Nanri A, Mizoue T, Takahashi Y, Noda M, Inoue M, Tsugane S; Japan Public Health Center-based Prospective Study Group. Weight change and all-cause, cancer and cardiovascular disease mortality in Japanese men and women: the Japan Public Health Center-Based Prospective Study. Int J Obes (Lond). 2010;34:348–356.

14. Mulligan AA, Lentjes MAH, Luben RN, Wareham NJ, Khaw KT. Weight change and 15 year mortality: results from the European Prospective Investigation into Cancer in Norfolk (EPIC-Norfolk) cohort study. Eur J Epidemiol. 2018 Jan;33(1):37–53.

15. Stevens J Erber E Truesdale KP et al. Long- and short-term weight change and incident coronary heart disease and ischemic stroke: The Atherosclerosis Risk in Communities Study Am J Epidemiol 2013 178 2 239-248

16. Chei CL, Iso H, Yamagishi K, et al. Body mass index and weight change since 20 years of age and risk of coronary heart disease among Japanese: the Japan Public Health Center-Based Study, Int J Obes (Lond), 2008, vol. 32 1(pg. 144–151)

17. Taubman SL, Robins JM, Mittleman MA, Hernán MA. Intervening on risk factors for coronary heart disease: an application of the parametric g-formula. Int J Epidemiol. 2009;38:1599–1611

18. Danaei G, Robins JM, Young JG, Hu FB, Manson JE, Hernán MA. Weight Loss and Coronary Heart Disease: Sensitivity Analysis for Unmeasured Confounding by Undiagnosed Disease. Epidemiology. 2016 Mar;27(2):302–10.

19. Wannamethee SG, Shaper AG, Walker M. Overweight and obesity and weight change in middle aged men: impact on cardiovascular disease and diabetes. J Epidemiol Community Health. 2005;59:134–139.

20. Douglas IJ, Bhaskaran K, Batterham RL, Smeeth L Bariatric Surgery in the United Kingdom: A Cohort Study of Weight Loss and Clinical Outcomes in Routine Clinical Care. PLoS Med 2015; 12(12): e1001925

21. Yarnell JW, Patterson CC, Thomas HF, Sweetnam PM. Comparison of weight in middle age, weight at 18 years, and weight change between, in predicting subsequent 14-year mortality and coronary events: Caerphilly Prospective Study. J Epidemiol Community Health. 2000; 54: 344–348.

22. Koudstaal S P-RM, Denaxas S, Gho Jmih, Shah AD, Gale CP, Hoes AW, Cleland JG, Asselbergs FW, Hemingway H. Prognostic burden of heart failure diagnosed in primary care or secondary care: a population-based linked electronic health record cohort study in 2.1 million people. Eur J Heart Fail. 2017;19(9):1119–1127

23. Denaxas S, Gonzalez-Izquierdo A, Direk K, Fitzpatrick N, Fatemifar G, Banerjee A, Dobson R, Howe L, Kuan V, Lumbers T, Pasea L, Patel R, Shah A, Hingorani A, Sudlow C, and Hemingway H. UK phenomics platform for developing and validating electronic health record phenotypes: CALIBER. JAMIA 2019: doi: 10.1093/jamia/ocz105

24. Kim ES, Jeong JS, Han K et al.Impact of weight changes on the incidence of diabetes mellitus: a Korean nationwide cohort study. Sci Rep. 2018;8(1):3735. doi: 10.1038/s41598-018-21550-3

25. Hernán MA, Taubman SL. Does obesity shorten life? The importance of well-defined interventions to answer causal questions. 3:S8–14. doi: 10.1038/ijo.2008.82.

26. Hernán, Miguel A., and VanderWeele, Tyler J. 2011. Compound treatments and transportability of causal inference. Epidemiology 22(3): 368–77

27. Force USPST, Curry SJ, Krist AH, Owens DK, Barry MJ, Caughey AB, et al. Behavioral weight loss interventions to prevent obesity-related morbidity and mortality in adults: US Preventive Services Task Force recommendation statement. JAMA 2018;320(11):1163–71

28. Meschia JF, Bushnell C, Boden-Albala B, Braun LT, Bravata DM, Chaturvedi S, et al. Guidelines for the primary prevention of stroke: a statement for healthcare professionals from the American Heart Association/American Stroke Association. Stroke 2014; 45:3754–3832

29. Dale CE, Fatemifar G, Palmer TM et al. Causal Associations of Adiposity and Body Fat Distribution With Coronary Heart Disease, Stroke Subtypes, and Type 2 Diabetes Mellitus: A Mendelian Randomization Analysis.Circulation. 2017 Jun 13;135(24):2373–2388

