## Appendix for "Weight change and the incidence of cardiovascular diseases in adults with normal weight, overweight and obesity without chronic diseases; emulating trials using electronic health records"

### Section 1: Assumptions for variables – Calculation of baseline date

We calculated theoretical weight and weight change measurements every year after the 1st measurement of bodyweight for each individual. We assumed linear trend and we censored observations of participants with >4 years without weight measurement.

The formula for the linear trend will be

$$w(t_{hyp}) = \frac{365.25 * [w(t+1) - w(t)]}{x} + w(t)$$

where $w(t_{hyp})$ indicates the (hypothetical) bodyweight measurement at time $t_{hyp}$, calculated from previous and next measurements w(t) and w(t+1) respectively and x indicates the number of days between dates of actual measurements t and t+1.

Below, we present a hypothetical participant, who measured her weight 4 times

1st bodyweight measurement at 30/6/99

2nd bodyweight measurement at 7/1/01

3rd bodyweight measurement at 25/4/02

4th bodyweight measurement at 4/7/07

For this individual, we assumed that she had the 1st bodyweight measurement at 30/6/99 was 84.8kg (this will be the baseline measurement). The 2nd (hypothetical this time) measurement will be at 30/6/00, 80.5kg and the other hypothetical measurement is 79.5 kg at 30/6/01. These 3 bodyweight measurement are represented in orange colour below. We censored the last observation from this individual, as her last measurement was >4 years after 25/4/02, so we did not assume linear trend between these 2 observations.

So the 1st weight change measurement will be (84.8 – 80.5)/ 80.5 ≈ –5.1%

Her 2nd weight change measurement will be (80.5– 79.5)/ 79.5≈ –1.2%

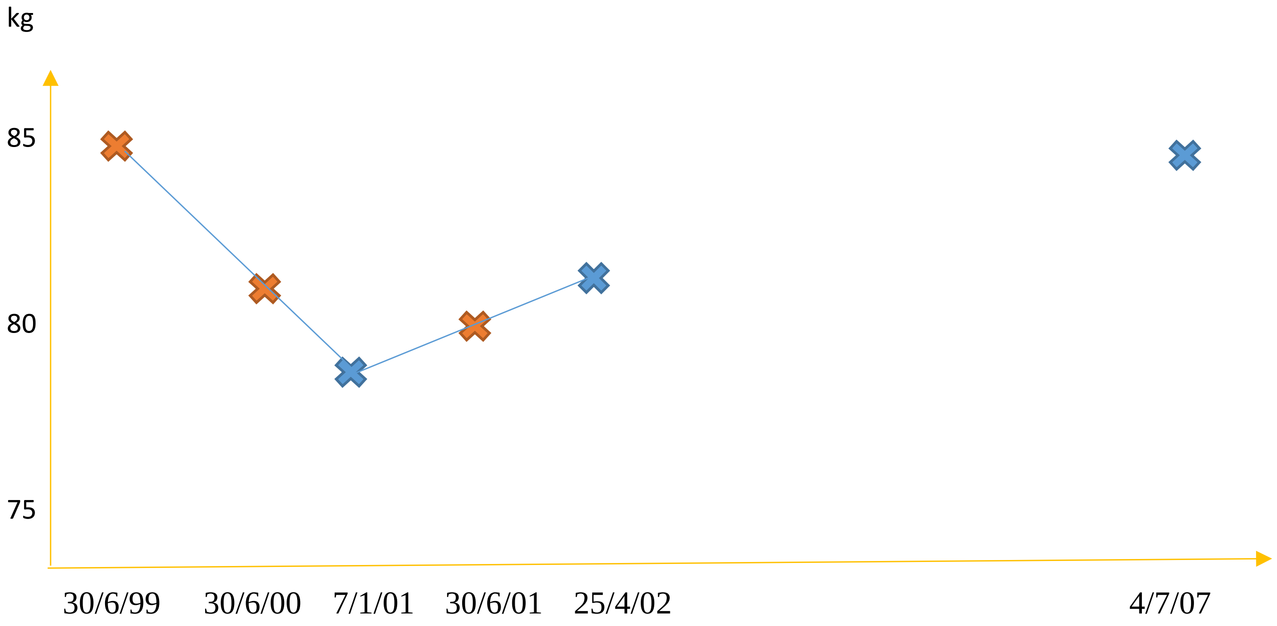

##### *Calculation of smoking status, physical activity and baseline date*

We considered smoking status (in the main analysis) and physical activity (in the sensitivity analysis) as confounders in our study. We assumed that their last observation carries forward for 4 years, unless the participant has another measurement of these variables. We implemented complete-case analysis, requiring that all individuals should have observations of smoking status and physical activity (in the sensitivity analysis). For this reason, we created time "windows" within which could be the baseline of our hypothetical interventions.

The date of baseline for an individual was defined as the date of the first BMI and weight observations, in which we would have valid information on smoking status. For those individuals with information on both physical activity and smoking status, we selected as baseline date, the date of the first BMI and weight observations, in which they had information on both smoking status and physical activity.

Below, we present a hypothetical individual who had her 1st bodyweight measurement at 20/5/2003 and her 2nd at 30/6/2006 (she also had a few more after 2006). Given that she had her 1st smoking status record at 2004 and her 1st physical activity observation at 2005, we considered that the time "window" for her potential baseline would be between 7/1/2005 and 30/6/2008. So, her actual baseline in this example would be at 30/6/2006 (time of her 2nd bodyweight measurement).

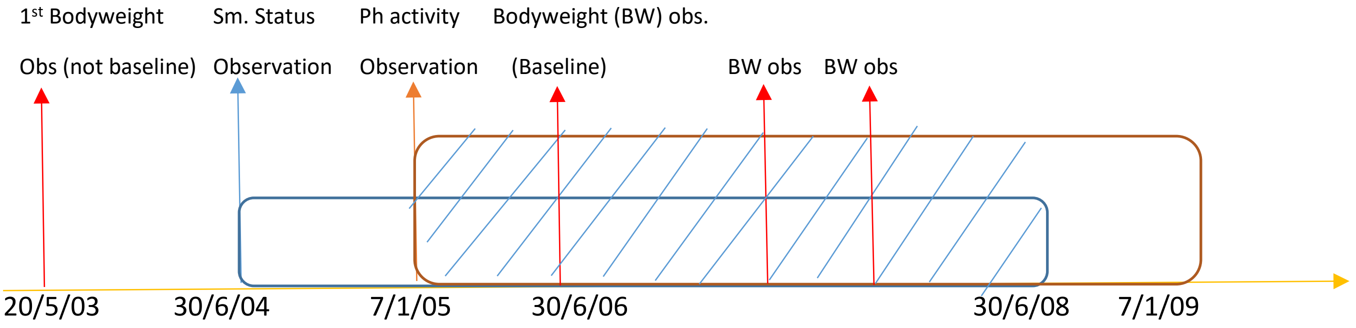

### Section 2: Protocol of the target trials – protocol of the emulated trials

***PROTOCOL OF THE TARGET TRIALS***

***Eligibility criteria***

The trials would enrol healthy men and non-pregnant women aged 45-69 years old, from primary care practices in England registered between 1998 and 2016 who had not undergone bariatric surgery in the past and without any prevalent cardiovascular disease, diabetes, cancer (apart from non-melanoma skin cancer), severe mental diseases (acute stress, phobia, anxiety, schizophrenia, depression, bipolar disorder or affective disorder), major inflammatory diseases (systemic lupus erythematosus, rheumatoid arthritis, gout and ulcerative colitis), Parkinson's disease, multiple sclerosis, renal disease and renal failure at baseline. Moreover, individuals who either die or develop one of the aforementioned chronic diseases or the outcome or women who become pregnant during the 1$^{st}$ year of the hypothetical interventions, would be excluded from the trials. The trials would be conducted separately among normal weight, overweight and obese individuals (see Table 1 and Figure 1). We would additionally consider these trials, stratified by sex (males and females) and age (<60 & ≥60 years old).

***Treatment strategies***

Individuals in the trials would be randomly assigned to the following two-year hypothetical weight change interventions

a) lose ≥3% & <20% of their weight each year or undergo bariatric surgery

b) maintain their weight, defined as weight change >-3% & <3% of bodyweight each year

c) gain ≥3% & <20% of their weight each year.

Under all three strategies, individuals would be allowed to deviate from their assigned weight change intervention after 2 years. From these trials, we considered women who became pregnant and individuals who developed chronic diseases during the 2nd year of follow up as a clinically allowable reason for deviating from assigned intervention; thus, these individuals will be allowed to deviate from their assigned intervention in the 2nd year. The chronic diseases considered as clinically allowable conditions to deviate from the intervention in these trials would be the following; diabetes, cancer (apart from non-melanoma skin cancer), dementia, severe mental diseases (acute stress, phobia, anxiety, depression, schizophrenia, bipolar disorder and affective disorder), chronic kidney disease, chronic obstructive pulmonary disease, HIV, major inflammatory diseases(systemic lupus erythematosus, rheumatoid arthritis, gout, and inflammatory bowel disease), Parkinson's disease, multiple sclerosis and renal failure. When diabetes or non-melanoma skin cancer were the outcomes of interest (positive and negative control outcomes), we also considered CVD as a clinically allowable condition for deviating from the initial intervention. For more details, see table 1.

***Treatment assignment***

Individuals would be randomly assigned to a treatment strategy at baseline

***Follow-up***

Follow-up would start at randomization and would end at diagnosis of a cardiovascular event, death, loss to follow-up, 7 years after baseline, or 31st June 2016, whichever occurs first.

***Endpoints***

Primary endpoint;

Composite CVD outcome (CVD death, non-fatal Myocardial infarction, non-fatal stroke, hospitalisation from coronary heart disease and heart failure)

Secondary endpoints;

a) Composite CHD outcome (CHD death, non-fatal Myocardial infarction, hospitalisation from coronary heart disease)

b) fatal and non-fatal Myocardial infarction,

c) fatal and non-fatal stroke,

d) heart failure

e) CVD deaths

f) Diabetes – used as a positive control outcome (i.e. it is expected that weight loss reduces and weight gain increases the risk of diabetes)

g) Non melanoma skin cancer – used as a negative control outcome (i.e. it is expected that no effect of weight loss or gain on non-melanoma skin cancer)

***Causal contrasts***

Per-protocol effect, defined as the effect of adhering to the assigned intervention for two years

***Analysis plan***

*i. Data structure*: Follow-up would be divided into one year periods in which the weight change intervention per year was recorded, along with information on confounders, CVD outcome (1:yes , 0:no), death (1:yes , 0:no) and loss to follow-up (1:yes , 0:no) recorded during each year.

*ii. Outcome regression:* Pooled logistic regression model would be used to estimate the hazard ratios of the hypothetical interventions and the cumulative incidence risk curves of each intervention.

*iii. Emulating randomisation at baseline:* Individuals would be randomised at baseline *iv. Dealing with non-adherence to intervention:.* Inverse probability of treatment weights (IPTW) would be used to adjust for time-fixed and time-dependent confounders. In IPTW, each observation would be weighted by the inverse of the probability of an individual having received his/her observed weight change intervention during the 2$^{nd}$ year, given his/her past intervention and prognostic factors history. To calculate the denominator of the IPTW, we would use multinomial logistic regression models for the observed weight change intervention (i.e. weight loss, maintenance and gain) during the 2$^{nd}$ year, in which we would include the determinants of the observed intervention at the 2$^{nd}$ year, i.e. prognostic factors measured before baseline, during the 1$^{st}$ year (as described above) and during the 2$^{nd}$ year of these interventions, along with the observed weight change intervention during the 1$^{st}$ year. The IPTW remained constant after the second year, because we would be interested in the effect of a weight change strategy sustained over 2 years only. After calculating the IPTW for the received intervention in the second year, individuals would be then censored during the second year, if they deviated from their assigned intervention. For those individuals who would develop a chronic disease other than CVD during the 2$^{nd}$ year (and thus were free to deviate in the 2$^{nd}$ year as well from their assigned intervention), they would be assigned the weight of 1 across all time points. We remark that we would use the non-stabilised weights, because the regime of the trials was dynamic (i.e. we had specified clinically allowable reasons after which individuals were free to deviate from their initial strategy)

*v. Dealing with loss to follow-up:* Additional adjustment for pre- and post-randomisation prognostic factors of loss to follow-up would also be used through inverse probability of censoring weighting (IPCW), to estimate the effect of the interventions, had the participants remained uncensored during the follow-up. These analyses would be thus valid under missing at random, given the covariates used to model the censoring mechanism.

*vi. Final calculation of IP weights*: IPTW and IPCW would be multiplied at each time point. The final weight for each individual for a specific time would be taken as the product of his/her weights up until that time point. We would truncate weights >15 (which were higher than the 99$^{th}$ percentile of weights) and would set it to 15.

*vii. Risk curves*: We would additionally estimate absolute risks for CVD, CHD and diabetes by fitting the pooled logistic models that were mentioned above, including product terms between treatment and follow-up time (time, squared time and cubic time) to allow for time varying effects. The estimated parameters would be then used to calculate the cumulative incidence of CVD, CHD and diabetes (see details in the Appendix – section 3).

*viii. Variance estimators:* We would use robust variance estimators to calculate 95% CI for the hazard ratio estimates, and we would use non-parametric bootstrapping from 500 samples to obtain percentile-based 95% CI for the cumulative incidence estimates.

#### *PROTOCOL OF THE EMULATED TRIALS*

##### *Eligibility criteria*

Same as for the target trials plus we additionally excluded individuals with an excess number of bodyweight measurements or clinical consultations in the 1$^{st}$ year (defined as ≥6 bodyweight measurements or with ≥12 primary care consultations), under the assumption that these individuals would be too unhealthy to participate in the study.

##### *Treatment strategies*

Same as for the target trials plus we excused individuals from following their assigned intervention if they have ≥12 clinical consultations or measured their bodyweight ≥6 times per year in the primary care[‡] during the 2$^{nd}$ year.

##### *Treatment assignment*

Patients are classified into one of three weight change groups (maintenance, loss, or gain) based on their observed weight status. Randomization is emulated via adjustment for baseline covariates.

***Follow-up***

Same as for the target trials

***Endpoints***

Same as for the target trials

***Causal contrasts***

Observational analog of the per-protocol effects

***Analysis plan***

Same as for the target trial, the only difference is that individuals are not randomised at enrolment. To emulate randomization at baseline, we adjusted for: age (in years), sex (men/ women), BMI (in kg/m$^2$), prevalence of hypertension (yes/no), record of high LDL levels (before baseline; yes/no), use of diuretics (before baseline; yes/no); family history of CVD (yes/no); hypertension (during the 1st year, yes/no); high LDL levels (during the 1st year; yes/no), use of diuretics (during the 1st year); smoking status (during the 1st year; never/ former/ current), number of weight measurements (during the 1st year; 1 meas./ 2 meas./ 3-5 meas.), number of clinical consultations (during the 1st year; 1-2 consultations, 3-5 consultations, 6-8 consultations, 9-11 consultations) and region (in categories; London/ South West/ South Central/ South East/ East/ West Midlands/ Central North and North West)

### Section 3: Details on the statistical analysis

##### *Disease models*

To estimate the hazard ratios in all analyses, we used pooled logistic regression models. In these models, we use the following notation; *t* is the follow-up time (in years), $A_{t,wl}$ denotes the value of the "weight loss intervention" between t and t+1, $A_{t,wm}$ denotes the value of the "weight maintenance intervention" between t and t+1, $A_{0,wg}$ and $A_{t,wg}$ indicates the value of the "weight gain intervention" between t and t+1, $\boldsymbol{L_t}$ indicates a vector of covariates (number of weight measurements [categorical; 1: 1 meas, 2:2 meas., 3: 3-5 meas), number of clinical consultations (ordered; 1: 1-2 clinical cons., 2: 3-5 clinical cons., 3: 6-8 clinical cons, 4: 9-11 clinical cons), record of hypertension, record of high LDL measurement, use of diuretics] measured between t and t+1. Of note, $\boldsymbol{L_0}$ (i.e. baseline confounders) additionally contains information on age (at baseline), sex, region, BMI (in kg/m$^2$ at baseline), prevalence of hypertension (at baseline – criteria; systolic blood pressure>140mmol/Hg or diastolic blood pressure>90mmol/Hg), record of high LDL measurement (before baseline), use of diuretics (before baseline), apart from the information of number of weight measurements, number of clinical consultations, record of hypertension, high LDL measurement and use of diuretics that occurred during the 1$^{st}$ year (i.e. between t=0 and t=1). Moreover, $D_t$ indicates the values of the CVD outcome between t and t+1 and $C_t$ denotes whether an individual was censored between t and t+1. The over-bars represent the previous history of a variable from the beginning of follow-up and the superscript *T* indicates a transpose of a vector of parameters. Throughout our analyses, we censored the person-time when someone discontinued his/her initial treatment assignment, because we were interested in the per-protocol effect of our "interventions". That is, we fit the model:

$$logit\left[pr\left(D_{t+1}=1\middle|A_0,L_0,\overline{D}_t=0,\overline{C}_{t+1}=0\right)\right]=\beta_{0,t}+\beta_1A_{0,wl}+\beta_2A_{0,wg}+\ {\beta_3}^T\boldsymbol{L_0} \quad \text{(S.1)}$$

where $\overline{C}_{t+1}$ is the indicator for (artificial) censoring at time $t$+1, $\beta_{0,t}$ is a time-varying intercept, calculated as a constant plus a linear, a quadratic and a cubic term for time t. Moreover, $\beta_1$ and $\beta_2$ corresponds to the log hazard ratios for the weight loss and weight gain "interventions" respectively (compared to the weight maintenance "intervention").

***Models for IP weights***

We were interested in the per-protocol effect of our "trails", so we weighted our disease models with the inverse probability of treatment weights (IPTW). The IPTW correspond to the reciprocal of the probability that an individual adhered his/her observed weight change intervention given his past treatment and pre and post baseline prognostic factors history. We used the unstabilised version of IPTW because the regime of our trials was dynamic (if people were not healthy at year 1, they were free to deviate from their allocated intervention).

During the 1$^{st}$ year, individuals adhered to the initial intervention they were allocated (by default in our observational analog of the target trial), so the IPTW during the 1$^{st}$ year was 1 for all individuals (i.e. the probability that an individual adhered his/her observed weight change intervention was 1, the same for the inverse of this probability). For the 2nd year (i.e. year 1), we calculated the IPTW as follows.

The unstabilized inverse probability weights for each patient at year 1 (i.e. the 2nd year) are defined as

$$IPTW_1 = \frac{1}{f(A_1|A_0, L_0, L_1, \overline{D}_1 = 0\ )}$$

As described elsewhere, (2, 3) we fit the multinomial logistic model

$$logit\left[pr\left(A_1 = j \middle| A_0, L_0, L_1, \overline{D}_1 = 0\right)\right] = \varphi_0 + \varphi_1 A_0 + \varphi_2^T L_0 + \varphi_3^T L_1 \qquad \text{(S.2)}$$

to estimate $f(A_1|A_0, L_0, L_1, \overline{D}_1 = 0\ )$, where j=1,2,3 is the "intervention" of interest.

To be consistent with the notation, we used previously in the outcome regression model,

if $A_1 = 1 \Rightarrow A_{1,wl} = 1, A_{1,wm} = 0, A_{1,wg} = 0$

if $A_1 = 2 \Rightarrow A_{1,wl} = 0, A_{1,wm} = 1, A_{1,wg} = 0$

if $A_1 = 3 \Rightarrow A_{1,wl} = 0, A_{1,wm} = 0, A_{1,wg} = 1$

If the participants develop a severe disease at year 1, then the participants are free to deviate from their intervention. These individuals will not be used in model (S.2), i.e. only healthy individuals contribute in the calculation of the probability of adhering to their intervention in year 1 [$f(A_1|A_0, L_0, L_1, \overline{D}_1 = 0\ )$], and will have IPTW=1. Of note, we run this model before excluding the individuals who did not adhere to their allocated intervention during the 2nd year. Moreover, from year 2 onwards, participants were free to deviate from their hypothetical weight change intervention, so the weights remained constant till the end of their follow-up.

We also took into consideration the potential bias due to loss to follow-up, by calculating the effect of an intervention, had the participants remained uncensored. To implement that, we additionally multiply IPTW calculated from (S.2) with the reciprocal of the probability of remaining uncensored

More specifically, we calculate the inverse of the probability of remaining uncensored at each time point

$$IPCW_t = \prod_{k=0}^{t} \frac{1}{f(C_{k+1} = 0 | A_k, \overline{L_k}, \overline{C_k} = 0, \overline{D_{k+1}} = 0)}$$

by running the pooled logistic regression

$$logit[pr\ (C_{t+1} = 1 | A_t, \overline{L_t}, C_t = 0, \overline{D_{t+1}} = 0)] = d_{0,t} + d_1 A_{t,wl} + d_2 A_{t,wg} + \ d_3{}^T \boldsymbol{L_0} + \ d_4{}^T \boldsymbol{L_t} \quad \textbf{(S.4)}$$

to estimate $f(C_{k+1} = 1 | A_k, \overline{L_k}, \overline{C_k} = 0, \overline{D_{k+1}} = 0)$ at each time point. Then we calculate $f(C_{k+1} = 0 | A_k, \overline{L_k}, \overline{C_k} = 0, \overline{D_{k+1}} = 0)$ =1- $f(C_{k+1} = 1 | A_k, \overline{L_k}, \overline{C_k} = 0, \overline{D_{k+1}} = 0)$ at each time point and finally we multiply these probabilities through all time points, to estimate $\prod_{k=0}^{t} f(C_{k+1} = 0 | A_k, \overline{L_k}, \overline{C_k} = 0, \overline{D_{k+1}} = 0)$. Then the $IPCW_t$ is the reciprocal of this probability in each time point.

For IPCW, we used smoking status only for the 1st year (i.e. in the vector $L_0$). We didn't use information from smoking status in the vector $L_k$, with k≥1, from follow-up, due to near positivity violations.

Table S2: Example of the dataset from 3 hypothetical individuals;

| **ID** | **Date** | **Time (in years)** | **$BW^t$** Bodyweight measurement at t | **$WC^t = BW^{t+1} - BW^t$** Weight change between t and t+1 | **$A^t$** Hypothetical intervention during t (1=weight loss, 2=weight maint., 3=weight gain) | **$C^t$** Censored between t and t+1 | **$C^{t+1}$** Censored between t+1 and t+2 | **$D^t$** Develop the CVD outcome between t and t+1 | **$D^{t+1}$** Develop the CVD outcome between t+1 and t+2 | **$L^t$** (example) N of Weight measurements (Confounder measured between t and t+1) | Severe disease measured between t and t+1 (from table 1) | **IPTW** |
|---|---|---|---|---|---|---|---|---|---|---|---|---|
| 1 | 5/5/2002 | 0 | 70 | -0.05 | 1 | 0 | 0 | 0 | 0 | 1 | 0 | 1 |
| 1 | 5/5/2003 | 1 | 66.5 | -0.098 | 1 | 0 | 1 | 0 | 0 | 2 | 0 | w1 |
| ~~1~~ | ~~5/5/2004~~ | ~~2~~ | ~~60~~ | | | ~~1~~ | ~~.~~ | ~~0~~ | ~~0~~ | ~~1~~ | ~~0~~ | ~~w1~~ |
| 2 | 3/8/2001 | 0 | 50 | 0 | 2 | 0 | 0 | 0 | 0 | 1 | 0 | 1 |
| 2 | 3/8/2002 | 1 | 50 | 0.02 | 2 | 0 | 0 | 0 | 0 | 1 | 0 | w2 |
| 2 | 3/8/2003 | 2 | 51 | . | 2 | 0 | 0 | 0 | 0 | 2 | 0 | w2 |
| 2 | 3/8/2004 | 3 | . | . | 2 | 0 | 0 | 0 | 0 | 1 | 0 | w2 |
| 2 | 3/8/2005 | 4 | . | . | 2 | 0 | 0 | 0 | 0 | 2 | 0 | w2 |
| 2 | 3/8/2006 | 5 | . | . | 2 | 0 | 0 | 0 | 1 | 2 | 0 | w2 |
| ~~2~~ | ~~3/8/2007~~ | ~~6~~ | ~~.~~ | ~~.~~ | ~~2~~ | ~~0~~ | ~~0~~ | ~~1~~ | ~~.~~ | ~~1~~ | ~~0~~ | ~~w2~~ |
| 3 | 9/6/2006 | 0 | 100 | -0.1 | 1 | 0 | 0 | 0 | 0 | 2 | 0 | 1 |
| 3 | 9/6/2007 | 1 | 90 | 0.11 | 1 | 0 | 1 | 0 | 0 | 1 | 1 | 1 |
| ~~3~~ | ~~9/6/2008~~ | ~~2~~ | ~~100~~ | ~~.~~ | ~~1~~ | ~~1~~ | ~~.~~ | ~~0~~ | ~~0~~ | ~~1~~ | ~~1~~ | ~~1~~ |

*Participant 1 was censored at 7/8/2004, participant 2 developed CVD at 10/11/2007 and participant 3 was censored at 12/10/2008*

*Description of table S2*

Participant ID=1 was allocated in the weight loss arm (she lost ≥3% of her bodyweight at year 0 and year 1). She was censored at 7/8/2004, during the 3rd year, so this was recorded in her year 2 raw in $C^t$ and in her year 1 in $C^{t+1}$. We subsequently delete any observations from this participant from time 2 onwards (orange line).

Participant ID=2 was allocated in the weight maintenance arm (her bodyweight change was <3 % at year 0 and year 1). She remained in the weight maintenance arm from year 2 onwards, even if she has no other measures of bodyweight, as after the completion of 2 years, people are free to deviate from their intervention. She developed the CVD outcome during the 7th year (10/11/2007).

Participant ID=3 was allocated in the weight loss arm (she lost ≥3% of her bodyweight at year 0). She developed cancer (which is included in the set of the severe chronic diseases described in the paper, so she was free to deviate from her intervention) during the 2nd year of follow-up, at 11/12/2007. Even if at time 1 (i.e. during the second year) she gained weight, he remained in the weight reduction arm. Her IPTW is 1 (because she developed cancer at year 1). She was censored during her 3rd year, so we subsequently deleted any observations from this participant from time 2 onwards.

***Risk curves***

After calculating the IPTW from (S.2), we fitted a (weighted) pooled logistic regression model as the one described in the beginning of section 1 in (S.1), with the only difference being that we add product terms with a linear, a quadratic and a cubic term for time t in the variables for weight loss and weight gain "intervention" at baseline. More specifically, the model we fitted is

$$logit\left[pr\left(D_{t+1} = 1 \middle| A_0, L_0, \overline{D}_t = 0, \overline{C}_{t+1} = 0\right)\right] =$$

$$= c_{0,t} + c_1 A_{0,wl} + c_2 A_{0,wg} + c_3 A_{0,wl} * t + c_4 A_{0,wg} * t + c_5 A_{0,wl} * t^2 + c_6 A_{0,wg} * t^2 + c_7{}^T \boldsymbol{L_0} \qquad \text{(S.3)}$$

Then, we created a dataset with all the time points under each "treatment", by copying each subject 3 times (one copy for every arm). We predicted the probability of the events from the (S.3) at each time point t and then we calculated probabilities that the participants remained free from the CVDs [S(t)=1-pr(D(t)=1)] for each person each year under all 3 "interventions". We then multiplied these probabilities (i.e. of remaining free from the outcome) through time t. Finally, we averaged the adjusted time-to-event curves over all subjects, so we obtained marginal time-to-event curves under each "intervention". Finally, we calculate the risk at each time point, by subtracting the marginal time-to-event probabilities from one.

***Sensitivity analyses***

1) *Including physical activity, index of multiple deprivation or ethnicity*

We implement complete case analyses, requiring from all individuals to have measurements of physical activity or ethnicity at baseline. We assume in the calculation of the time “windows” of physical activity and smoking status that the last observation carries forward for at most 4 years.

2) *Impact of pre-clinical diseases*

To take into consideration the impact of potential bias due to preclinic diseases, we assume that a chronic disease occurred one, two or three years before it was recorded during the follow-up time and we check whether our results remain the same in this sensitivity analysis. The set of chronic diseases we considered for this sensitivity analysis was: diabetes, cancer (apart from non-melanoma skin cancer), dementia, severe mental diseases (acute stress, phobia, anxiety, depression, schizophrenia, bipolar disorder and affective disorder), chronic kidney disease, chronic obstructive pulmonary disease, HIV, major inflammatory diseases(systemic lupus erythematosus, rheumatoid arthritis, gout, and inflammatory bowel disease), Parkinson’s disease, multiple sclerosis and renal failure

### Section 4 – Extra results

Figure S1: Estimated hazard ratios for non-melanoma skin cancer (i.e. negative control outcome) under hypothetical weight change interventions, by BMI group and overall

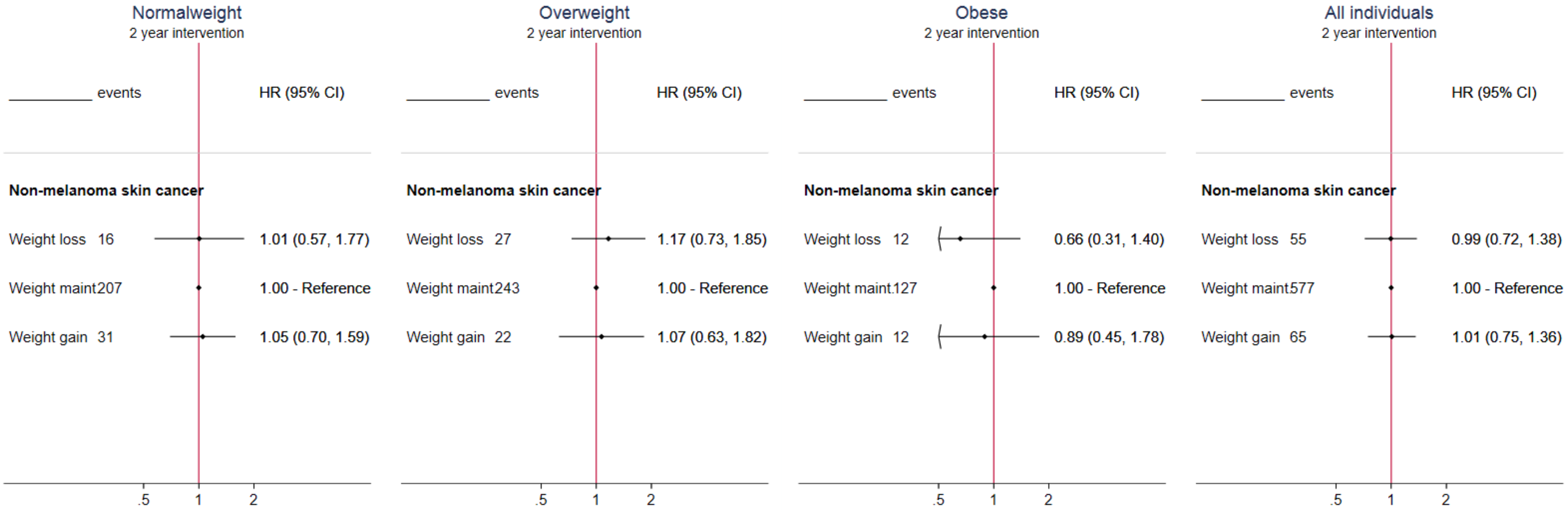

Figure S2: Percentage of individuals who developed a chronic disease (other than CVD) in the 2nd, 3rd and 4th year, by hypothetical interventions, BMI group

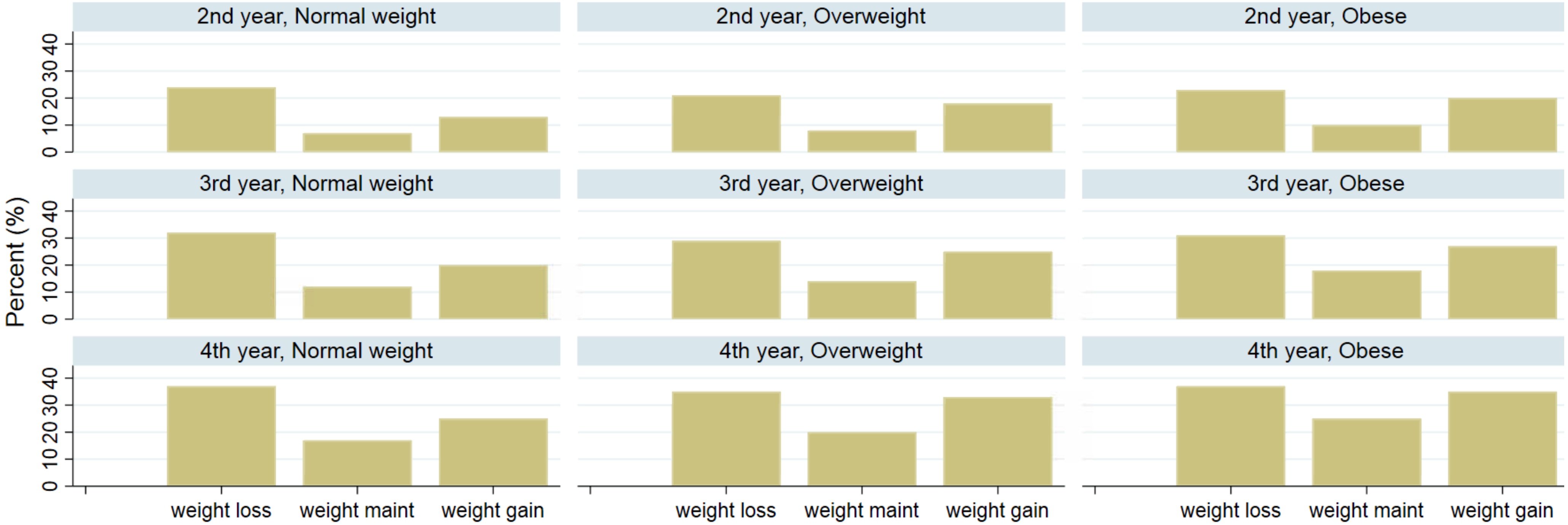

Figure S3: Hazard ratios of the per-protocol analysis emulated (two-year) interventions on cardiovascular diseases in normal weight individuals, using pooled logistic regression, by age group and sex

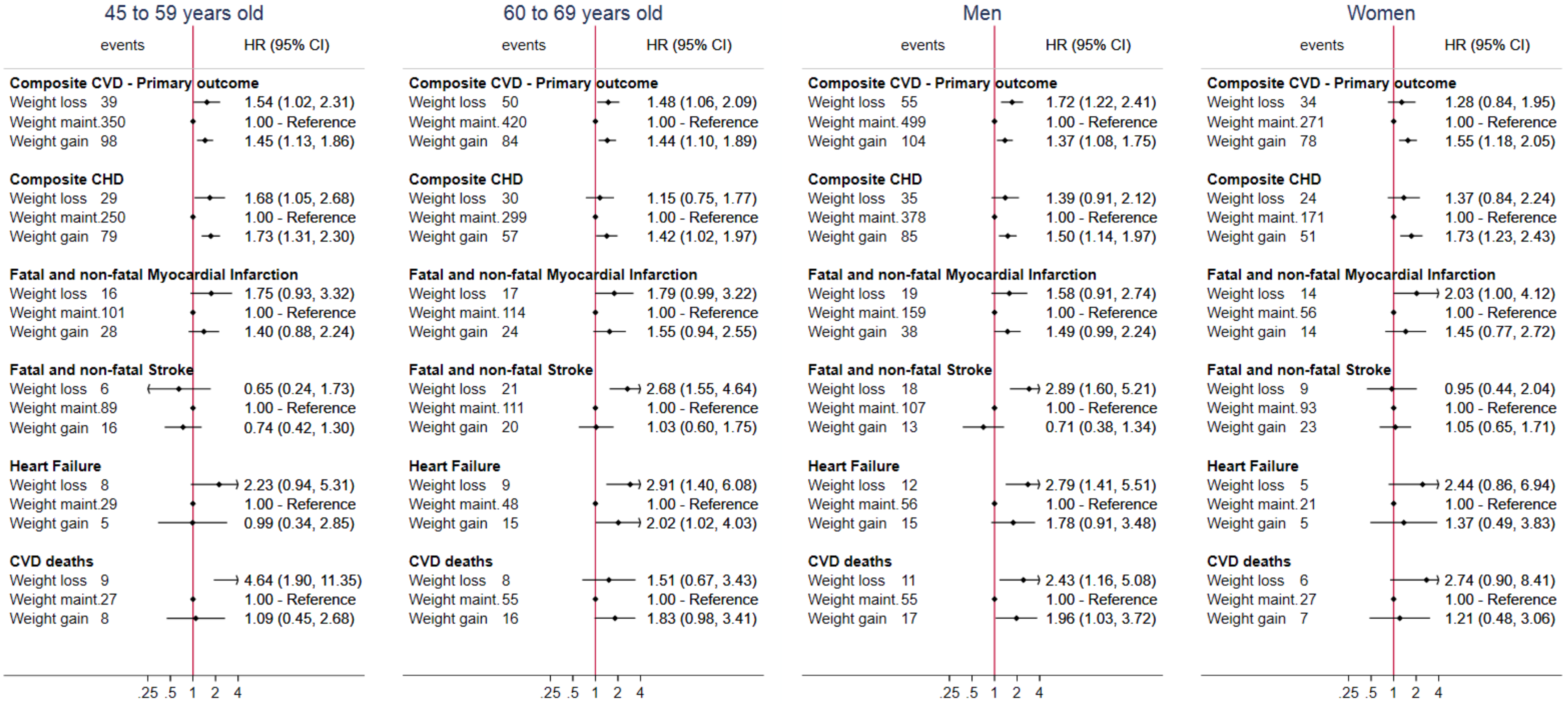

Figure S4; Hazard ratios of the per-protocol effect of the emulated interventions on the composite CVD outcome in normal weight, overweight and obese individuals. Sensitivity analysis includes further adjustment for index of multiple deprivation, ethnicity or physical activity

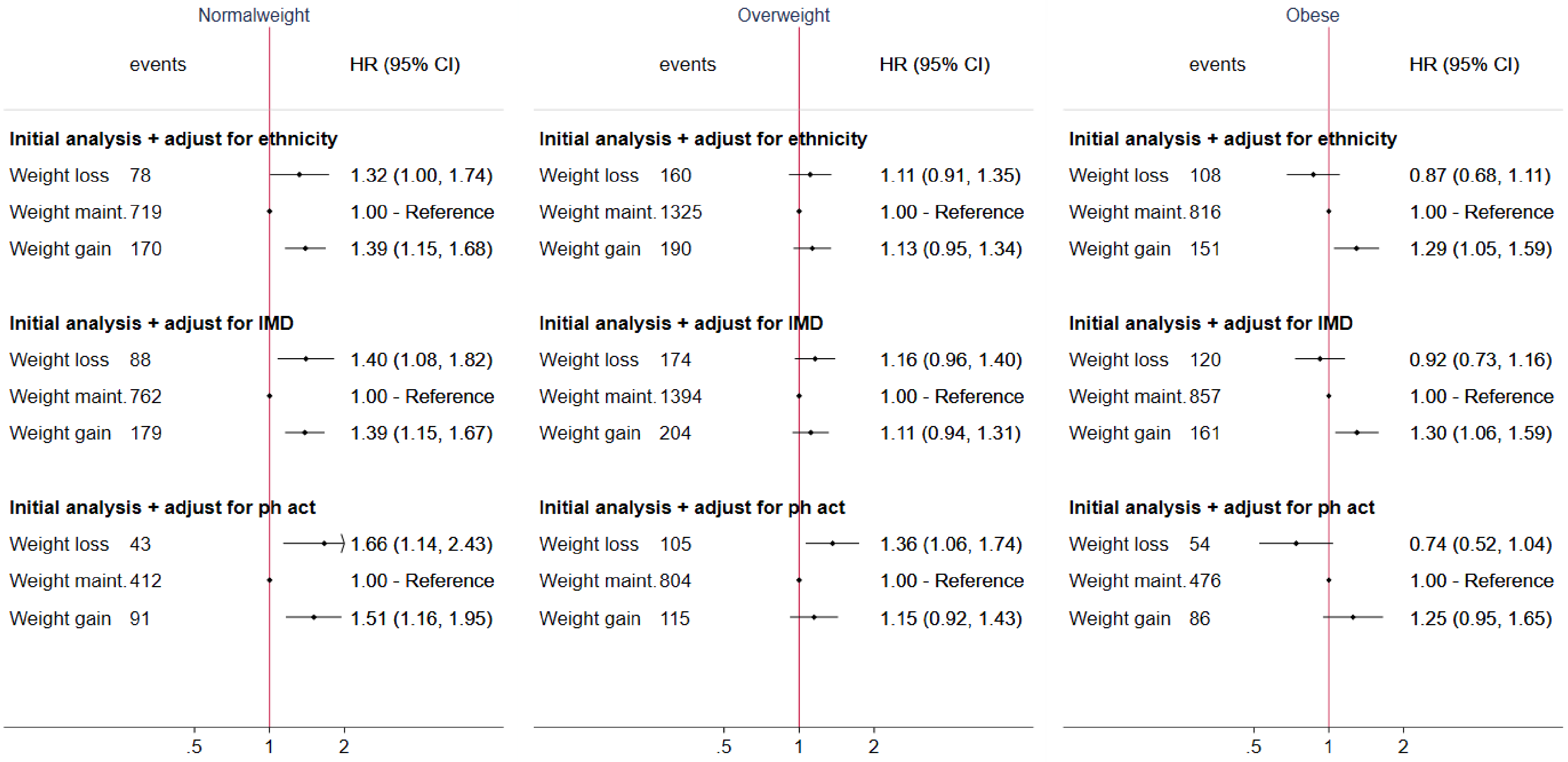

Figure S5; Hazard ratios of the emulated interventions on cardiovascular diseases in overweight individuals, using pooled logistic regression after assuming that a set of chronic diseases[1] occurred two years prior to the recorded date

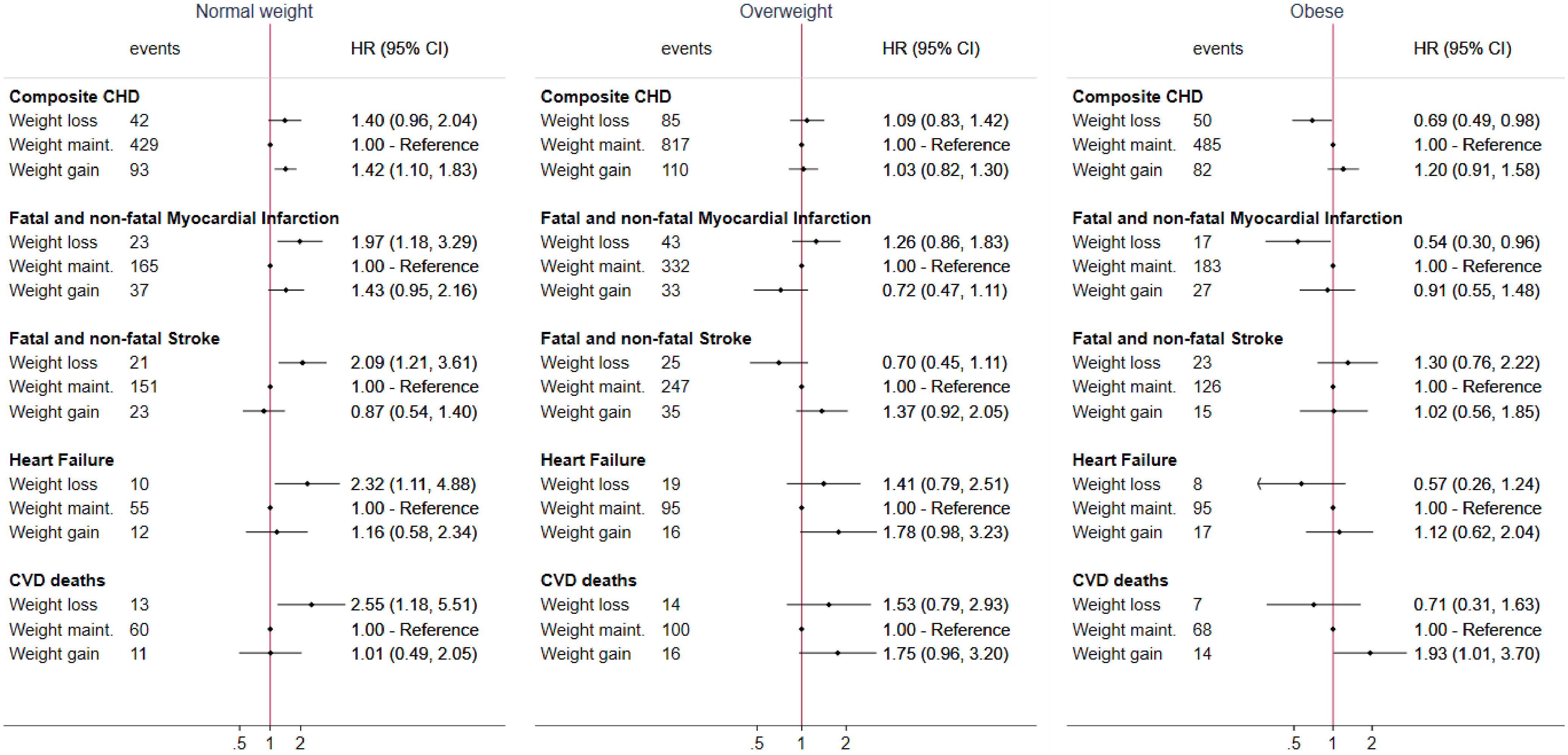


[1]Set of chronic diseases consists of: diabetes, cancer (apart from non-melanoma skin cancer), dementia, severe mental diseases (acute stress, phobia, anxiety, depression, schizophrenia, bipolar disorder and affective disorder), chronic kidney disease, chronic obstructive pulmonary disease, HIV, major inflammatory diseases(systemic lupus erythematosus, rheumatoid arthritis, gout, and inflammatory bowel disease), Parkinson's disease, multiple sclerosis and renal failure

Figure S6: Hazard ratios of the per-protocol analysis emulated (two-year) interventions on cardiovascular diseases in overweight individuals, using pooled logistic regression, by age group and sex

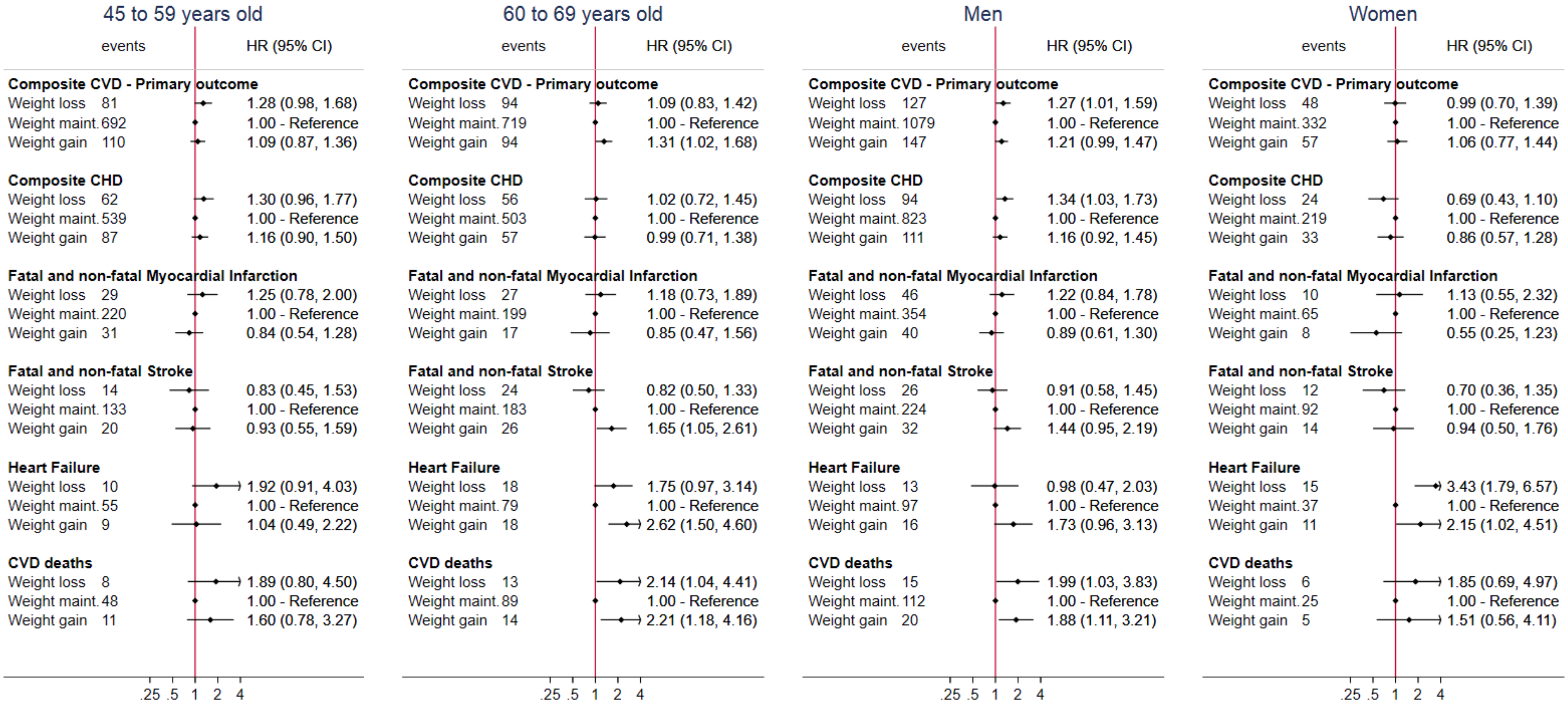

Figure S7: Hazard ratios of the per-protocol analysis emulated (two-year) interventions on cardiovascular diseases in obese individuals, using pooled logistic regression, by age group and sex

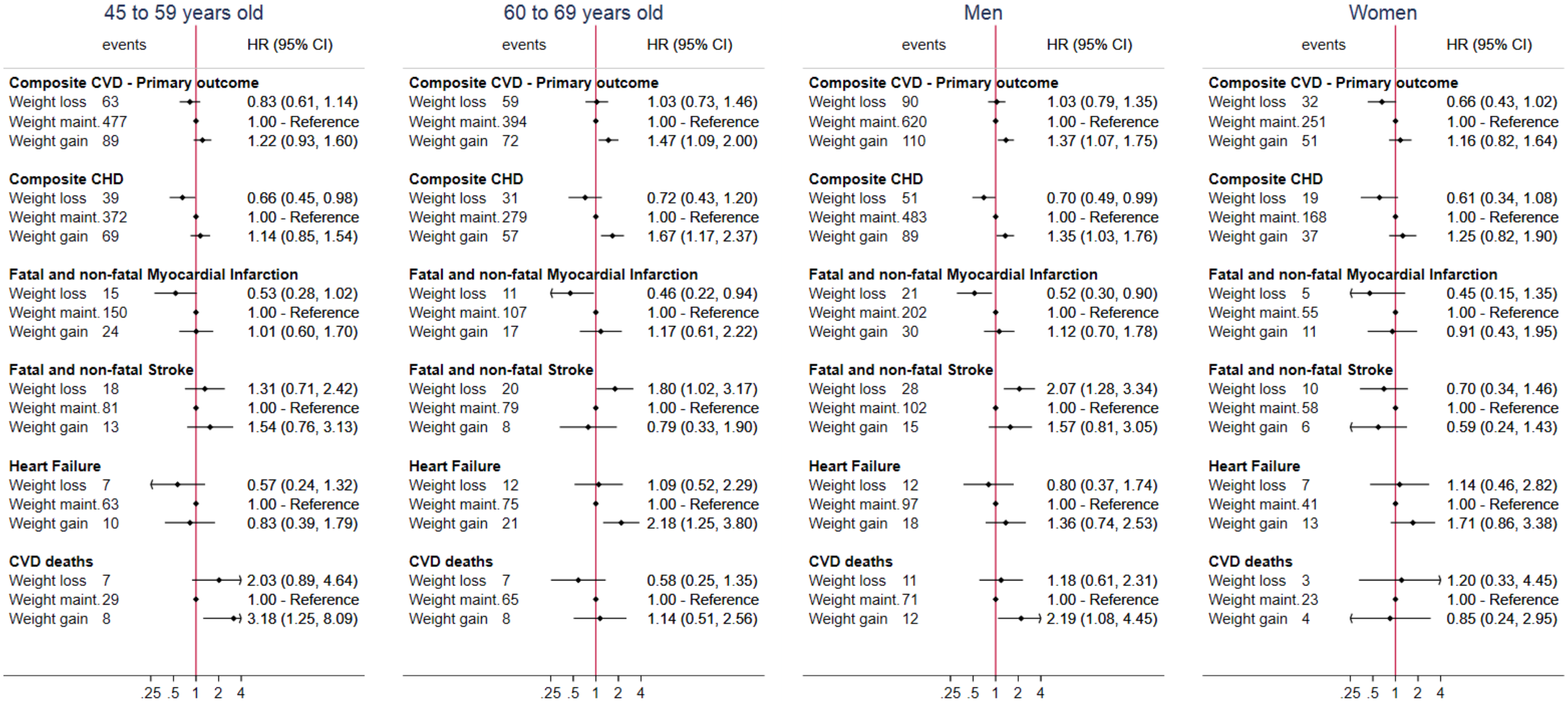

### Section 5 – Codelists used for the outcome definition

### 1) Primary outcome: Composite CVD

**The primary outcome (composite CVD) consists of**

**i. myocardial infarction,**
**ii. stable angina (hospitalization),**
**iii. unstable angina (hospitalization),**
**iv. other CHD (hospitalization),**
**v. heart failure (hospitalization),**
**vi. stroke**
**vii. CVD deaths**

**i. Myocardial infarction**

*a. Self-reported from primary care (CPRD)*
*b. ICD-10 codes from Hospital Episodes Statistics (HES)*

| ICD10 code | Interpretation |
|---|---|
| I252 | Old myocardial infarction |
| I21 | Acute myocardial infarction |
| I22 | Subsequent myocardial infarction |
| I23 | Certain current complications following acute myocardial infarction |
| I241 | Dressler's syndrome |

*c. OPSC codes from Hospital Episodes Statistics (HES)*

| OPSC code | Interpretation |
|---|---|
| K50.2 | Percutaneous transluminal coronary thrombolysis using streptokinase |
| K50.3 | Percutaneous transluminal injection of therapeutic substance into coronary artery NEC |

d. ICD-10 codes from ONS

| ONS ICD10 | Interpretation |
|---|---|
| I21 | Acute myocardial infarction |
| I22 | Subsequent myocardial infarction |
| I23 | Certain current complications following acute myocardial infarction |

e. ICD-9 codes from ONS

| ICD9 code | Interpretation |
|---|---|
| 410 | Acute myocardial infarction |
| 4110 | Other acute and subacute forms of ischemic heart disease ; Postmyocardial infarction syndrome |
| 4297 | Ill-defined descriptions and complications of heart disease ; Certain sequelae of myocardial infarction, not elsewhere classified |

#### ii. Stable Angina (hospitalization)

*a. ICD-10 codes from Hospital Episodes Statistics (HES)*

| ICD10 code | Interpretation |
|---|---|
| I201 | Angina pectoris with documented spasm |
| I208 | Other forms of angina pectoris |
| I209 | Angina pectoris, unspecified |

*b. OPSC codes from Hospital Episodes Statistics (HES)*

| OPCS code | Interpretation |
|---|---|
| K40 | Saphenous vein graft replacement of coronary artery |
| K40.1 | Saphenous vein graft replacement of one coronary artery |
| K40.2 | Saphenous vein graft replacement of two coronary arteries |
| K40.3 | Saphenous vein graft replacement of three coronary arteries |
| K40.4 | Saphenous vein graft replacement of four or more coronary arteries |
| K40.8 | Other specified saphenous vein graft replacement of coronary artery |
| K40.9 | Unspecified saphenous vein graft replacement of coronary artery |
| K41 | Other autograft replacement of coronary artery |
| K41.1 | Autograft replacement of one coronary artery NEC |
| K41.2 | Autograft replacement of two coronary arteries NEC |
| K41.3 | Autograft replacement of three coronary arteries NEC |
| K41.4 | Autograft replacement of four or more coronary arteries NEC |

| | |
|---|---|
| K41.8 | Other specified other autograft replacement of coronary artery |
| K41.9 | Unspecified other autograft replacement of coronary artery |
| K42 | Allograft replacement of coronary artery |
| K42.1 | Allograft replacement of one coronary artery |
| K42.2 | Allograft replacement of two coronary arteries |
| K42.3 | Allograft replacement of three coronary arteries |
| K42.4 | Allograft replacement of four or more coronary arteries |
| K42.8 | Other specified allograft replacement of coronary artery |
| K42.9 | Unspecified allograft replacement of coronary artery |
| K43 | Prosthetic replacement of coronary artery |
| K43.1 | Prosthetic replacement of one coronary artery |
| K43.2 | Prosthetic replacement of two coronary arteries |
| K43.3 | Prosthetic replacement of three coronary arteries |
| K43.4 | Prosthetic replacement of four or more coronary arteries |
| K43.8 | Other specified prosthetic replacement of coronary artery |
| K43.9 | Unspecified prosthetic replacement of coronary artery |
| K44 | Other replacement of coronary artery |
| K44.1 | Replacement of coronary arteries using multiple methods |
| K44.8 | Other specified other replacement of coronary artery |
| K44.9 | Unspecified other replacement of coronary artery |
| K45 | Connection of thoracic artery to coronary artery |
| K45.1 | Double anastomosis of mammary arteries to coronary arteries |
| K45.2 | Double anastomosis of thoracic arteries to coronary arteries NEC |
| K45.3 | Anastomosis of mammary artery to left anterior descending coronary artery |
| K45.4 | Anastomosis of mammary artery to coronary artery NEC |
| K45.5 | Anastomosis of thoracic artery to coronary artery NEC |
| K45.8 | Other specified connection of thoracic artery to coronary artery |
| K45.9 | Unspecified connection of thoracic artery to coronary artery |
| K46 | Other bypass of coronary artery |
| K46.1 | Double implantation of mammary arteries into heart |
| K46.2 | Double implantation of thoracic arteries into heart NEC |
| K46.3 | Implantation of mammary artery into heart NEC |
| K46.4 | Implantation of thoracic artery into heart NEC |
| K46.8 | Other specified other bypass of coronary artery |

| | |
|---|---|
| K46.9 | Unspecified other bypass of coronary artery |
| K44.2 | Revision of replacement of coronary artery |
| K45.6 | Revision of connection of thoracic artery to coronary artery |
| K46.5 | Revision of implantation of thoracic artery into heart |

##### iii. Unstable Angina (hospitalization)

*a. ICD-10 codes from Hospital Episodes Statistics (HES)*

| ICD10 code | Interpretation |
|---|---|
| I240 | Coronary thrombosis not resulting in myocardial infarction |
| I248 | Other forms of acute ischaemic heart disease |
| I249 | Acute ischaemic heart disease, unspecified |
| I200 | Unstable angina |

##### iv. Other CHD (hospitalization)

*a. ICD-10 codes from Hospital Episodes Statistics (HES)*

| ICD10 code | Interpretation |
|---|---|
| I250 | Atherosclerotic cardiovascular disease, so described |
| I251 | Atherosclerotic heart disease |
| I253 | Aneurysm of heart |
| I254 | Coronary artery aneurysm |
| I255 | Ischaemic cardiomyopathy |
| I256 | Silent myocardial ischaemia |
| I258 | Other forms of chronic ischaemic heart disease |
| I259 | Chronic ischaemic heart disease, unspecified |

*b. ICD-10 codes from ONS*

| ONS ICD10 | Interpretation |
|---|---|
| I20 | Angina pectoris |
| I21 | Acute myocardial infarction |
| I22 | Subsequent myocardial infarction |
| I23 | Certain current complications following acute myocardial infarction |
| I24 | Other acute ischaemic heart diseases |
| I25 | Chronic ischaemic heart disease |

*c. ICD-9 codes from ONS*

| ONS ICD9 | Interpretation |
|---|---|
| 410 | Acute myocardial infarction |
| 411 | Other acute and subacute forms of ischemic heart disease |
| 412 | Old myocardial infarction |
| 413 | Angina pectoris |
| 414 | Other forms of chronic ischemic heart disease |
| 4292 | Ill-defined descriptions and complications of heart disease ; Cardiovascular disease, unspecified |
| 4295 | Ill-defined descriptions and complications of heart disease ; Rupture of chordae tendineae |
| 4296 | Ill-defined descriptions and complications of heart disease ; Rupture of papillary muscle |
| 4297 | Ill-defined descriptions and complications of heart disease ; Certain sequelae of myocardial infarction, not elsewhere classified |

#### v. Heart failure (hospitalization)

##### *a. ICD-10 codes from Hospital Episodes Statistics (HES)*

| ICD10 | Interpretation |
|---|---|
| I50.0 | Congestive heart failure |
| I50.1 | Left ventricular heart failure |
| I50.9 | Heart failure, unspecified |
| I11.0 | Hypertensive heart disease with (congestive) heart failure |
| I13.0 | Hypertensive heart and renal disease with (congestive) heart failure |
| I32.2 | Hypertensive heart and renal disease with both (congestive) heart failure and renal failure |

##### *b. ICD-10 codes from ONS*

| ICD10 | Interpretation |
|---|---|
| I50.0 | Congestive heart failure |
| I50.1 | Left ventricular heart failure |
| I50.9 | Heart failure, unspecified |
| I11.0 | Hypertensive heart disease with (congestive) heart failure |
| I13.0 | Hypertensive heart and renal disease with (congestive) heart failure |
| I32.2 | Hypertensive heart and renal disease with both (congestive) heart failure and renal failure |

##### *c. ICD-9 codes from ONS*

| ICD9 | Interpretation |
|---|---|
| 428 | Congestive heart failure, unspecified |

| | |
|---|---|
| 428.1 | Left heart failure |
| 428.9 | Heart failure, unspecified |

#### vi. Stroke

*a. Self-reported from primary care (CPRD)*

*b. ICD-10 codes from Hospital Episodes Statistics (HES)*

| ICD10 code | Interpretation |
|---|---|
| I690 | Sequelae of subarachnoid haemorrhage |
| I61 | Intracerebral haemorrhage |
| I60 | Subarachnoid haemorrhage |
| I620 | Subdural haemorrhage (acute)(nontraumatic) |
| I621 | Nontraumatic extradural haemorrhage |
| I629 | Intracranial haemorrhage (nontraumatic), unspecified |
| I693 | Sequelae of cerebral infarction |
| I63 | Cerebral infarction |
| I690 | Sequelae of subarachnoid haemorrhage |
| I61 | Intracerebral haemorrhage |
| I60 | Subarachnoid haemorrhage |
| I620 | Subdural haemorrhage (acute)(nontraumatic) |
| I621 | Nontraumatic extradural haemorrhage |
| I629 | Intracranial haemorrhage (nontraumatic), unspecified |
| I691 | Sequelae of intracerebral haemorrhage |
| I692 | Sequelae of other nontraumatic intracranial haemorrhage |
| I694 | Sequelae of stroke, not specified as haemorrhage or infarction |
| I698 | Sequelae of other and unspecified cerebrovascular diseases |
| G463 | Brain stem stroke syndrome |
| G464 | Cerebellar stroke syndrome |
| G465 | Pure motor lacunar syndrome |
| G466 | Pure sensory lacunar syndrome |
| G467 | Other lacunar syndromes |
| I64 | Stroke, not specified as haemorrhage or infarction |

*c. ICD-10 codes from ONS*

| ONS ICD10 code | Interpretation |
|---|---|
| I690 | Sequelae of subarachnoid haemorrhage |
| I61 | Intracerebral haemorrhage |
| I60 | Subarachnoid haemorrhage |
| I620 | Subdural haemorrhage (acute)(nontraumatic) |
| I621 | Nontraumatic extradural haemorrhage |
| I629 | Intracranial haemorrhage (nontraumatic), unspecified |
| I693 | Sequelae of cerebral infarction |
| I63 | Cerebral infarction |
| I690 | Sequelae of subarachnoid haemorrhage |
| I61 | Intracerebral haemorrhage |
| I60 | Subarachnoid haemorrhage |
| I620 | Subdural haemorrhage (acute)(nontraumatic) |
| I621 | Nontraumatic extradural haemorrhage |
| I629 | Intracranial haemorrhage (nontraumatic), unspecified |
| I694 | Sequelae of stroke, not specified as haemorrhage or infarction |
| I698 | Sequelae of other and unspecified cerebrovascular diseases |
| I64 | Stroke, not specified as haemorrhage or infarction |
| I672 | Cerebral atherosclerosis |
| I679 | Cerebrovascular disease, unspecified |

*d. ICD-9 codes from ONS*

| ICD9 code | Interpretation |
| --- | --- |
| 431 | Intracerebral hemorrhage |
| 430 | Subarachnoid hemorrhage |
| 4321 | Other and unspecified intracranial hemorrhage ; Subdural hemorrhage |
| 4320 | Other and unspecified intracranial hemorrhage ; Nontraumatic extradural hemorrhage |
| 4329 | Other and unspecified intracranial hemorrhage ; Unspecified intracranial hemorrhage |
| 433 | Occlusion and stenosis of precerebral arteries |
| 434 | Occlusion of cerebral arteries |
| 431 | Intracerebral hemorrhage |
| 430 | Subarachnoid hemorrhage |
| 4321 | Other and unspecified intracranial hemorrhage ; Subdural hemorrhage |
| 4320 | Other and unspecified intracranial hemorrhage ; Nontraumatic extradural hemorrhage |
| 4329 | Other and unspecified intracranial hemorrhage ; Unspecified intracranial hemorrhage |
| 436 | Acute, but ill-defined, cerebrovascular disease |
| 4370 | Other and ill-defined cerebrovascular disease ; Cerebral atherosclerosis |
| 4379 | Other and ill-defined cerebrovascular disease ; Unspecified |

##### vii. CVD deaths

*a. ICD-10 codes from ONS: All “I” codes*

#### 2) Secondary outcome: Composite CHD

**Composite CHD consists of**

**i. myocardial infarction,**
**ii. stable angina (hospitalization),**
**iii. unstable angina (hospitalization),**
**iv. other CHD (hospitalization),**
